# Gain and loss of function variants in EZH1 disrupt neurogenesis timing and cause overlapping neurodevelopmental disorders

**DOI:** 10.1101/2022.08.09.22278430

**Authors:** Carolina Gracia-Diaz, Yijing Zhou, Qian Yang, Chul-Hwan Lee, Paula Espana-Bonilla, Shuo Zhang, Natàlia Padilla, Raquel Fueyo, Garrett Otrimski, Dong Li, Sarah Sheppard, Paul Mark, Margaret H. Harr, Hakon Hakonarson, Lance Rodan, Adam Jackson, Pradeep Vasudevan, Corrina Powel, Shehla Mohammed, Sateesh Maddirevula, Hamad Alzaidan, Eissa A. Faqeih, Stephanie Efthymiou, Valentina Turchetti, Fatima Rahman, Shazia Maqbool, Vincenzo Salpietro, Shahnaz H Ibrahim, Gabriella di Rosa, Henry Houlden, Conchi Estaras, Anna C.E. Hurst, Michelle L. Thompson, Anna Chassevent, Constance L. Smith-Hicks, Xavier de la Cruz, Alexander Holtz, Erin Torti, MJ Hajianpour, Claudine Rieubland, Dominique Braun, Siddharth Banka, Genomic England Research Consortium, Elizabeth A. Heller, Murielle Saade, Hongjun Song, Guo-Li Ming, Fowzan S. Alkuraya, Reza Maroofian, Pankaj B. Agrawal, Danny Reinberg, Elizabeth J. Bhoj, Marian A. Martinez-Balbas, Naiara Akizu

**Author notes:** These authors contributed equally (Carolina Gracia-Diaz, Yijing Zhou).

## Abstract

Genetic disruption of chromatin regulators is frequently found in neurodevelopmental disorders (NDDs). While chromatin regulators are attractive therapeutic targets, studies to determine their implication in the etiology of NDDs are limited, preventing advances in diagnosis and treatment strategies. Here, we uncover pathogenic variants in the chromatin modifier *Enhancer of Zeste Homologue 1 (EZH1)* as the cause of overlapping recessive and dominant NDDs in 17 individuals. *EZH1* encodes one of the two alternative histone H3 lysine 27 (K27) methyltransferases of the Polycomb Repressive Complex 2 (PRC2). Unlike the other PRC2 subunits, which are associated with the pathogenesis of human cancers and developmental disorders, the implication of EZH1 in human development and disease is largely unknown. Using cellular models and biochemical studies, we demonstrate that recessive variants cause EZH1 loss of function (LOF), while dominant variants are all missense mutations that modify the catalytic activity of EZH1, thus generating a gain of function (GOF) effect. Consistent with a pathogenic effect, depletion or overexpression of EZH1 perturbs neuronal differentiation in the developing chick embryo neural tube. Furthermore, using human pluripotent stem cell (hPSC) derived neural cultures and forebrain organoids, we show that *EZH1* LOF and GOF variants respectively delay and accelerate the schedule of cortical projection neuron generation. Our work identifies *EZH1* LOF and GOF variants as the genetic basis of previously undefined NDDs and uncovers an essential role of EZH1 in regulating the timing of neurogenesis.

## Introduction

Neurodevelopmental disorders (NDDs) are a group of conditions that arise from impaired development of the central nervous system and include developmental delay and intellectual disability among the most severe manifestations. To date, hundreds of genes, predominantly affected by *de novo* mutations, have been implicated in the etiology of NDDs^1–3^. Approximately half of these mutations are nonsense, frameshift or splice site variants that cause loss of function, while the remainder are missense mutations with often unpredictable molecular consequences that are rarely interrogated at a functional level^3, 4^. Thus, despite recent advances in next generation sequencing and molecular diagnosis, many variants remain pathogenically undefined, contributing to the diagnostic odyssey of the affected families and lack of treatment options for patients^5^.

A group of NDD genes particularly susceptible to pharmacological intervention are chromatin regulators, especially those encoding subunits of Polycomb Repressive Complex 2 (PRC2) ^6–10^. The core PRC2 complex is formed by EED, SUZ12, and one of the two catalytic subunits, EZH1 or EZH2, that sequentially mono-, di- and tri-methylate the lysine at position 27 of the histone H3 (H3K27me1, me2 and me3 respectively). Through the establishment and propagation of H3K27me3, PRC2 maintains transcriptional repression of genes that regulate cellular identities during development and tissue homeostasis in the adult^11^. The involvement of PRC2 subunits and H3K27me3 demethylases, KDM6A and KDM6B, in the etiology and prognosis of cancers, has motivated an upsurge of pharmacological inhibitors with therapeutic potential ^12, 13^. In addition, dominant *de novo* pathogenic variants in *EZH2*, *EED* and *SUZ12* are a common cause of Weaver and Cohen Gibson Syndromes which are characterized by overgrowth and intellectual disability^8, 14, 15^, and KDM6B and KDM6A are implicated in autism and Kabuki Syndrome^16–18^. Notably, the current availability of H3K27 methyltransferase and demethylase inhibitors renders these H3K27me3 associated developmental disorders susceptible for therapeutical targeting.

Despite evidence involving PRC2 in human development and disease, the implication of EZH1 is less recognized. EZH1 has traditionally been considered the minor catalytic subunit of PRC2, in part because its histone lysine methyltransferase activity (HMT) is weaker than that of EZH2^19^. During development, EZH2 catalyzes most of the H3K27me3 that represses differentiation genes. Thus, upon deletion of EZH2, EZH1 can only partially maintain H3K27me3 and transcriptional repression^20–22^. Consequently, *Ezh2* deletion is fatal for embryonic progression^23^. In contrast, *Ezh1* deletion barely affects H3K27me3 maintenance at early developmental stages and is compatible with life, at least in mouse and zebrafish^24, 25^. Nevertheless, a growing body of evidence suggests that EZH1 is important during differentiation and for postmitotic cells. For instance, EZH1 is required for myogenic gene activation during skeletal muscle differentiation, for atrophy induced response of myotubes and neonatal heart regeneration in mice ^26–28, 29^. In addition, EZH1 represses multipotency of hematopoietic stem cells in favor of more lineage restricted embryonic progenitors^30^ and a switch from EZH2 to EZH1 expression co-occurs with the onset of blood cell differentiation^31^. Indeed, the reduction of EZH2 and maintenance or increase of EZH1 expression during differentiation is common to most cell types, but its functional relevance remains poorly understood.

Similarly, little is known about the contribution of EZH1 to brain development and function. Although *Ezh1* knock out mice do not show overt cellular and molecular brain defects, *Ezh1* protects neurons from degeneration upon *Ezh2* deletion^32^. Moreover, *Ezh1* is required for synaptic development of cultured hippocampal neurons^33^. Here, we show that *EZH1* is necessary for human brain development and function. Using next generation sequencing methods, we identified biallelic truncating and heterozygous missense *EZH1* variants in 17 individuals with varying degrees of developmental, language and motor delays, mild to severe intellectual disability, dysmorphic facial features. Notably, the lack of overgrowth as a hallmark distinguishes these patients from those with PRC2-EZH2 associated developmental syndromes. In biochemical studies and cellular models, we demonstrate that patients’ *EZH1* variants cause recessive LOF or dominant GOF *EZH1* effects. Furthermore, through functional studies in the chick embryo neural tube development model and in genetically edited human pluripotent stem cells (hPSC) derived cortical neuron and forebrain organoids, we establish that EZH1 LOF and GOF delay and accelerate neuronal differentiation respectively. Together, our work shows that a precise regulation of EZH1 activity is required to coordinate neurogenesis timing and uncovers EZH1 LOF and GOF variants as the genetic basis of previously undefined overlapping NDDs.

## Results

### Identification of EZH1 variants in individuals with undefined neurodevelopmental delay

As part of our ongoing efforts to provide genetic diagnosis for patients with undefined neurodevelopmental disorders, we performed exome and genome sequencing on a child showing cognitive, speech and motor development delay. The co-occurrence of the neurodevelopmental symptoms with an atypical facial dysmorphism suggested a genetic cause for the disease. Results pointed to a heterozygous missense variant in *EZH1 (*NM_001991.5: *c.2203C>G;* p.L735F) as the strongest candidate. Through collaborations facilitated by Genematcher^34^, Deciphering Developmental Disorders project^35^, and 100,000 Genomes Project^36^ we expanded our cohort to 17 individuals from 13 unrelated families, all sharing a clinical phenotype of neurodevelopmental delay and carrying distinct variants in *EZH1* (Fig 1, Supplementary Table 1), which supports the implication of *EZH1* in the disease pathogenesis.

**Figure 1:**
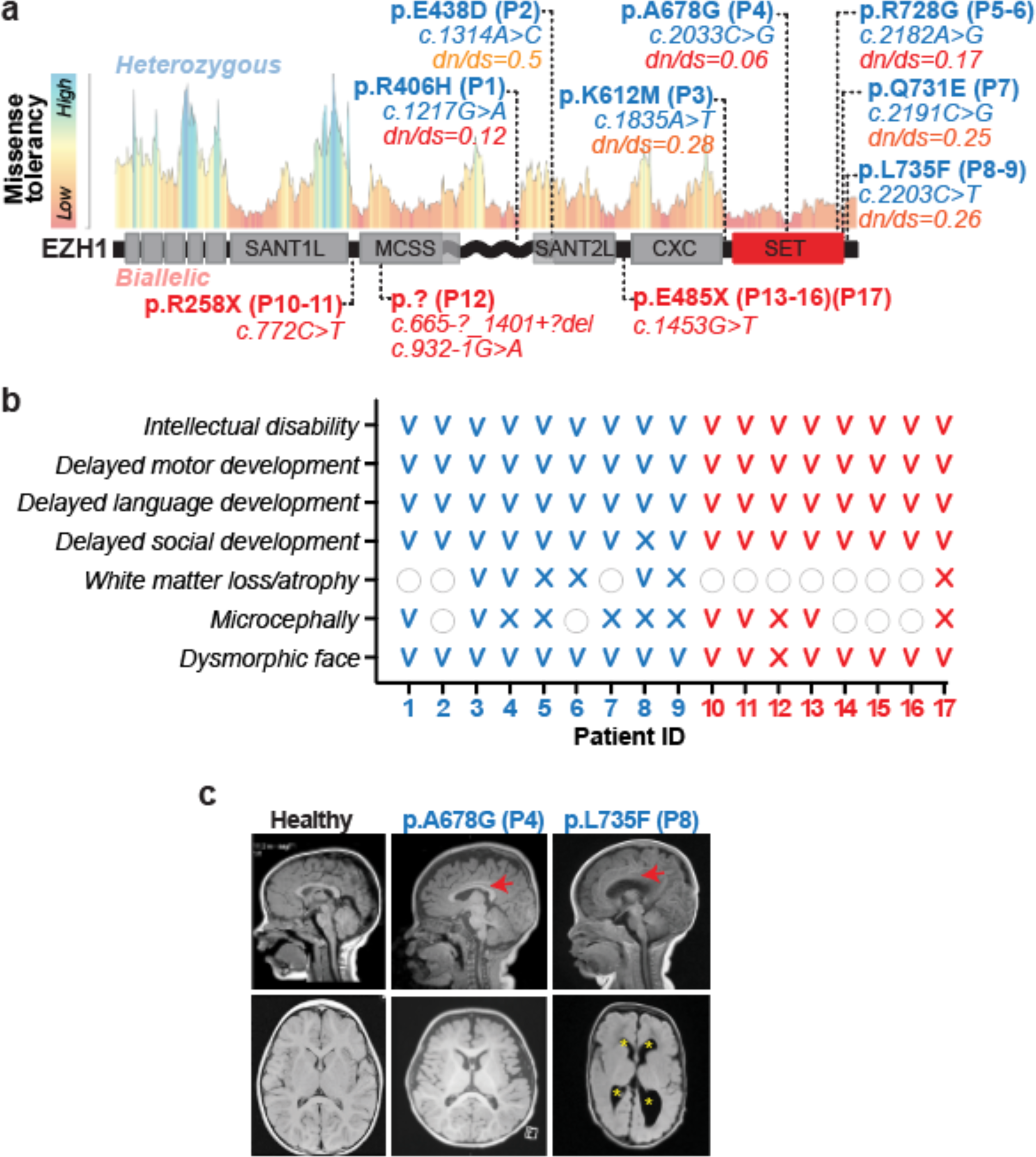
Heterozygous and biallelic variants in *EZH1* are associated with previously undiagnosed neurodevelopmental disorders. **a,** Schematic representation of EZH1 protein and its domains with the missense tolerance landscape from MetaDome. The localizations of EZH1 variants and the corresponding patient IDs are indicated. Heterozygous variants are labeled in blue with their missense tolerance score is indicated. Biallelic variants are labeled in red and depicted below the EZH1 schematic representation. **b,** Summary of major clinical features in patients with heterozygous variants (blue) and biallelic variants (red). V=present, X=absent, empty circle=unknown. **c**, Brain MRIs, sagittal (up) and axial (down), of two patients with the indicated heterozygous variant. Red arrow indicates thinner corpus callosum. Yellow asterisks indicate enlarged ventricles.

Further supporting the pathogenicity of these variants, *EZH1* is a gene with low missense variant rate in the general population (*Z*=4.2) and loss of function variants are rare (o/e=0.26). In addition, our patients’ variants are absent in over 140,000 sequences of a sample population reported in GnomAD v2.1.1^37^. Among the variants identified, four are biallelic truncating variants present in eight individuals (Fig 1a). Two of the individuals are siblings of a consanguineous family carrying a homozygous nonsense (*c.772C>T;* p.R258X) variant inherited from their parents (Fig 2a). Another consanguineous family with four affected siblings carries a nonsense variant in homozygosity (*c.1453C>T*; p.E485X). Interestingly, an additional child from an unrelated consanguineous family of the same country carries the same homozygous variant and shares haplotype in chr17:40519338-41245021, suggesting a common founder event in the region. Sanger sequencing in available family members confirmed segregation of EZH1 variants according to a strict recessive mode of inheritance with full penetrance in these families (Fig 2b- c). The eighth child is the only affected of her family and carries two variants in compound heterozygosity: a large deletion covering exon 8-12 of *EZH1* inherited from the one parent (*c.6657-?_1401+?del*; p.?) and a *de novo* splice variant affecting the exon 10 splice acceptor site (c.932-1G>A; p.?) (Fig 1a, Fig 2d and Supplementary Fig 1a-b). RNA extraction from patient’s cells and analysis by RT-PCR confirmed exclusion of exon 10 (Fig 2d), which predicts a frameshift that introduces a premature stop codon in *EZH1* likely undergoing nonsense mediated decay. Accordingly, RT-qPCR of patient cells showed ∼80% reduction of total *EZH1* transcripts and Western Blot (WB) of cell lysates confirmed undetectable EZH1 levels (Fig 2e-f). This data indicates that biallelic *EZH1* variants cause loss of function (LOF).

**Figure 2:**
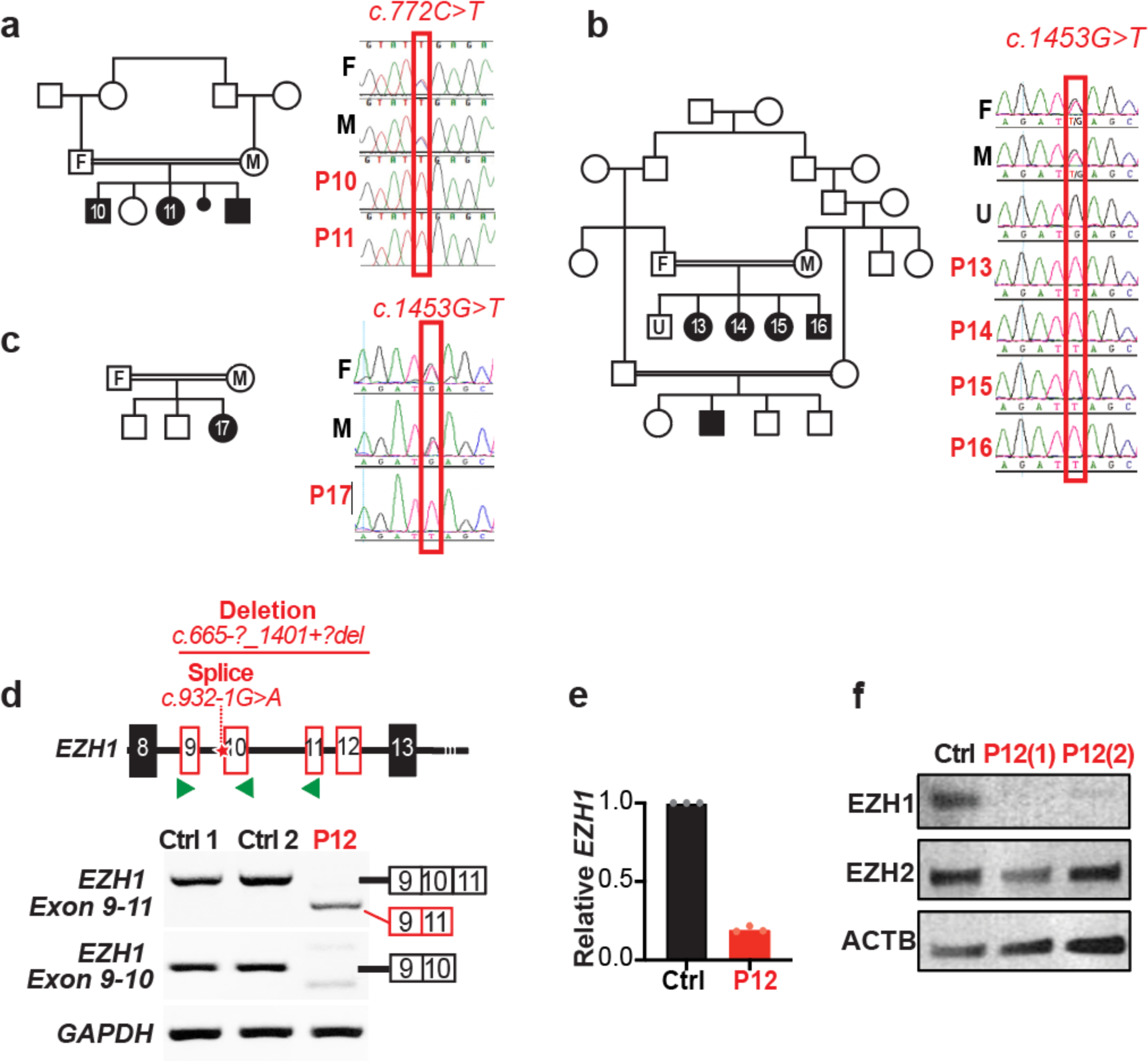
Biallelic EZH1 variants cause loss of function. **a-c,** Pedigree and sanger sequencing showing segregation of variants with the diseases in consanguineous families with **a,** 2 affected children (p10-11) harboring the homozygous nonsense *EZH1 c.772C>T*; p.R258X variant (note that the youngest sibling in the family was not included in the study) **b,** 4 affected children (P13-16) harboring the homozygous nonsense *EZH1* c.1453G>T: p.E485X variant. and **c,** a child (P17) from an unrelated family with shared haplotype and *EZH1* c.1453G>T: p.E485X variant. “F” indicates parent F, “M” parent M and “U” unaffected family members. **d,** Schematic representation of EZH1 exon 8-13, indicating the location of the deletion and splice variants (up). Green arrowheads indicate the location of the primers used for RT-PCR. RT-PCR of lymphoblastoid cell lines derived from two unrelated healthy individuals and the patient carrying the biallelic deletion and splice *EZH1* variants (P12) showing undetectable exon 10 containing transcripts in P12. *GAPDH* is shown as loading control. **e,** RT-qPCR analysis showing reduced *EZH1* levels in P12 lymphoblastoid cell lines compared to an unrelated control. Graph shows mean +/- SD of 3 technical replicates. **f,** Western blot analysis showing undetectable EZH1 levels and intact EZH2 in two independent P12 cell lysates compared to an unrelated control. ActinB is shown as loading control.

The remaining 9 patients carry heterozygous missense variants that affect evolutionarily conserved amino acids with low missense tolerance (Fig 1a, Supplementary Fig 2a). p.L735F and p.R728G are each present in two patients from unrelated families and the other five variants are unique to the affected individuals of five unrelated families. Parental DNA was obtained from three families and absence of the pathogenic variants in the parents of two (for p.A678G and p.R728G) confirmed *de novo* germline origin. In the third family, the variant (p.K612M) was detected at low frequency in one parent suggesting *de novo* somatic mosaicism with germline transmission to the affected child.

To estimate the effect of the missense variants in EZH1 function we analyzed them through molecular modeling. Four variants fall within or near the catalytic SET domain, which conserves 94% similarity with EZH2-SET domain^11^. Although the 3D structure of EZH1 has been recently resolved^38^, two of our patient variants fall within a gap in that structure. Thus, we used the 3D structure of EZH2-SET (PDB:5hyn^39^) as a template to model EZH1-SET domain and predict the effects of the new variants. Results for p.A678G, p.L735F and p.R728G show structural changes that may affect the kinetics of the methyltransferase reaction through different mechanisms (Supplementary Fig 2b). For example, the substitution of A678 residue for a glycine enlarges the H3K27 interaction pocket of EZH1 providing more flexibility to the structure and likely changing the stability of interactions with H3K27me0-me3. On the other hand, the aromatic ring of the phenylalanine that substitutes L735 residue can generate a stacking interaction with the adenine group of the methyl donor molecule (S-adenosyl methionine (SAM)) that could affect the turnover rate of SAM, and consequently, the efficiency of the H3K27me0 to me3 transition. R728 is localized in the vicinity of the SAM binding site and close in sequence to the residues forming the binding pocket. We expect that its replacement by a glycine, a residue conferring more conformational flexibility, may induce a local structure destabilization with an impact on binding. Visual analysis of p.K612M variants, shows that the substitution to a methionine could disrupt the intra-monomeric interactions of the native lysine, thus locally destabilizing the structure of EZH1. Alternatively, the location of 612 residue in the surface of EZH1 suggests that the methionine could also interfere with the interaction of EZH1 with other proteins. For the p.Q731E variant we were unable to predict the pathogenic mechanism given that the localization of the residue and the free energy calculations suggest a minor impact in the structure.

The remaining two variants fall near the MCSS-SANT2L loop of EZH1. This region contributes to the nucleosome binding of PRC2:EZH1 and differs in sequence similarity, structure and function from the equivalent region in EZH2^38^. The recently resolved 3D structures of PRC2:EZH1 bound and unbound to the nucleosome (PDB:7KSO and 7KSR^38^) allowed us to model the p.E438D variant, but not p.R406H due to a gap in the 3D structure. Visual analysis of EZH1 with E438 or D438 shows a change in the orientation of the side chain of the aspartic acid compared to the glutamic acid, particularly in the nucleosome bound PRC2:EZH1 structure (Supplementary Fig 2b). Furthermore, the free energy calculations suggest that the aspartic acid may destabilize the PRC2:EZH1 complex or its interaction with the nucleosome. Together, these data suggest that missense variants likely impact EZH1 catalytic activity through different mechanisms that affect the interactions with H3K27me0-me3, SAM or the nucleosome.

### Patients show neurodevelopmental delay with diverse clinical presentations regardless of variant type and zygosity

The 17 patients in our cohort share a neurodevelopmental disorder manifested early in life as global motor, speech and cognitive delay leading to intellectual disability, usually non-progressive and co-occurring with dysmorphic facial features (Fig 1b and Supplementary Table1). Seven patients (P3-6, P8-9, P17) had magnetic resonance imaging (MRI) and all show mild or unremarkable findings. P4 has mild white matter loss with otherwise unremarkable MRI. P8’s MRI is amongst the most severe showing mild cortical atrophy and white matter loss (Fig 1c). P8 and P9, who carry the same heterozygous variant (p.L735F), have hypoplastic optic nerves/chiasma on MRI. They also share phenotypes that correlate with atypical body fat accumulation, including an unusual fat distribution in legs in P8 and obesity and liver steatosis in P9.

Other clinical findings are variable and do not correlate with zygosity or type of genetic variant carried. For example, P9 has a history of severe autism, that was first suspected at few months of age due to a poor eye contact and tracking. Autism spectrum disorder (ASD) is being investigated in P7 as well, but there are no concerns for ASD in other patients. Aggressive behavior is reported in P5-6, who carry the same heterozygous variant (p.R728G), and in P10-11 siblings harboring the homozygous p.R258X variant. In addition, P8 started showing aggressive and obsessive-compulsive behaviors after 7 years of age. P1 and P4, who carry heterozygous missense variants, and P12 with biallelic LOF variants have all short stature (<15%ile), but height is normal in P8 and P9. P5 and P9 are the only patients with macrocephaly (>97%ile), while P1, P3, P10-11 and P13 have microcephaly. Additional sporadic findings include cardiovascular abnormalities in three patients with heterozygous missense variants, hypopigmentation in two and a hyperpigmented patch in a patient with biallelic LOF variant. Four patients (P1, P2, P8 and P14) have musculoskeletal abnormalities, including scoliosis in P1 and P2, and skeletal muscle biopsy in P4 shows mild defects in fiber size and NADH staining intensity (Supplementary Table 1). Notably, only two patients (P5 and P9) show overgrowth, with the remainder exhibiting normal or short stature. Together, these clinical presentations suggest that biallelic LOF and heterozygous missense *EZH1* variants cause similar NDDs that are clearly distinguishable from overgrowth with intellectual disability syndromes associated with PRC2-EZH2 variants. Additionally, differences in clinical features or severity between patients does not correlate with the type or zygosity of EZH1 mutations.

### Missense EZH1 variants promote hypermethylation of H3K27

Clinical similarities between patients led us to predict that the effect of the heterozygous *EZH1* missense variants could be similar to that of LOF variants, perhaps through a dominant negative effect on the catalytic activity. Thus, considering that most missense variants cluster within or near the catalytic SET domain of EZH1, we chose three of these variants to stably express in a human neural stem cell line (ReNCells) and monitor their effects on EZH1 expression and H3K27me3 levels. After sorting transduced ReNCells for EGFP co-expression we analyzed protein and histone lysates by WB. Results showed similar protein levels for wild type EZH1 (EZH1wt) and the three variants (Fig 3a). But, unexpectedly, p.A678G and p.Q731E displayed increased overall H3K27me3 levels compared to EZH1wt expressing ReNCells (Fig 3b). Consistently, chromatin immunoprecipitation and sequencing (ChIPseq) using H3K27me3 antibodies in p.A678G and EZH1wt expressing ReNCells confirmed a genome-wide increase in the proportion of H3K27me3 levels within peaks (Fig 3c). Furthermore, WB analysis of patient P4 fibroblasts carrying p.A678G in heterozygosity also showed a trend of increased H3K27me3 levels in comparison to control fibroblasts (Fig 3d). Together, these data suggest that at least some missense mutations create gain of function (GOF) EZH1 variants with increased methyltransferase activity. In line with this evidence, we found that EZH1-p.A678G is equivalent to the cancer-associated EZH2-p.A677G variant, which modifies the substrate preference of EZH2 leading to a change in H3K27me0 to me3 kinetics that results in increased H3K27me3 levels^40, 41^. Notably, there are several cancer-associated somatic EZH2 missense mutations with demonstrated GOF activity that are being investigated for therapeutic targeting with EZH1/2 inhibitors^13, 42^.

**Figure 3:**
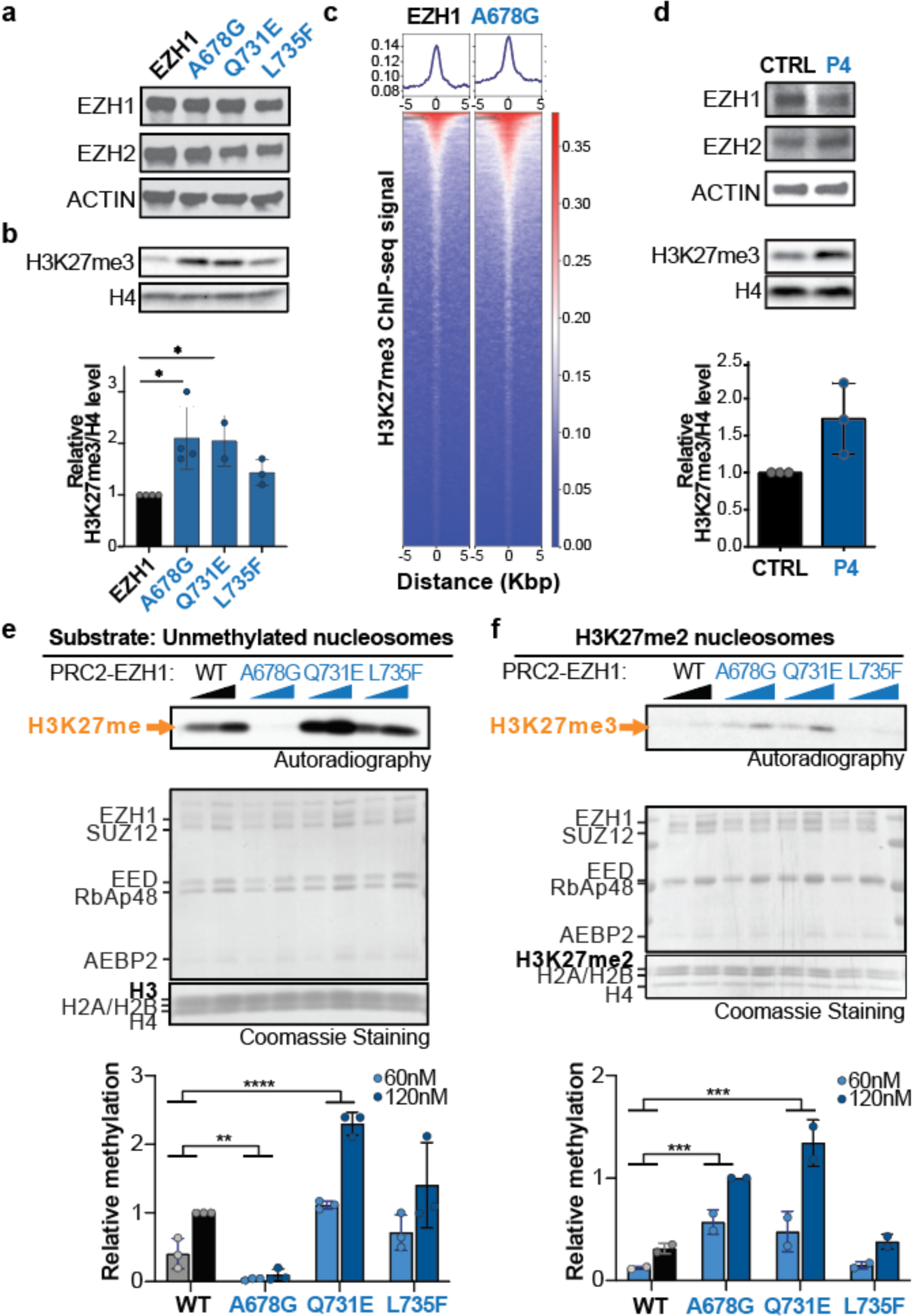
Heterozygous missense variants cause EZH1 gain of function that leads to hypermethylation of H3K27. **a-b,** Western blot analysis of EZH1, EZH2 (a) and H3K27me3 (b) levels from ReNcells transiently expressing either wild type or missense variants of EZH1. ActinB or H4 are shown as loading controls. Graph shows mean +/-SD of n=2-4 biological replicates. One-way ANOVA with Dunnett’s post hoc analysis test for multiple comparisons. *p- value<0.05. **c,** Enrichment plots showing average signal of H3K27me3 in ChIPseq peaks (top) and heatmaps showing normalized H3K27me3 ChIPseq intensities (bottom) ±5kb around the center of the peak. Plots represent data merged from 3 independent biological replicates. **d,** Western blot analysis of EZH1, EZH2 and H3K27me3 from P4 patient fibroblasts (top) and graph showing mean +/-SD of relative H3K27me3/H4 levels of n=3 independent fibroblasts lysates (bottom). **e-f,** Autoradiography and Coomassie stains of HMT assay reactions using two increasing concentrations of PRC2 complexes and unmethylated nucleosomes (e) or nucleosomes with dimethylated H3K27 (H3K27me2) as substrate (f). Graphs show mean +/-SD of n=2-3 independent assays. One-way ANOVA with Dunnett’s post hoc analysis test for multiple comparisons (combining mean of 60nM and 120nM assays). ***p-value<0.001, ****p- value<0.0001.

To corroborate if increased H3K27me3 levels by EZH1 missense variants is due to a molecular mechanism similar to cancer-associated EZH2 GOF variants, we performed *in vitro* histone methyltransferase (HMT) assays with reconstituted PRC2 complexes and assembled nucleosomes harboring predefined methylation states at H3K27. We first expressed the PRC2 subunits EED, SUZ12, RBAP48, AEBP2, together with EZH1wt or one of the three variants (p.A678G, p.Q731E or p.L735F) in a baculovirus expression system and intact complexes were successfully purified through tandem affinity purification for all (Fig 3e and f, Coomassie staining). Versions of PRC2 carrying EZH1wt or mutants were next incubated with tritiated methyl donor (SAM[^3^H]) and unmodified nucleosomes (H3K27me0) (Fig 3e) or nucleosomes harboring di- methylated H3K27 (H3K27me2) (Fig 3f) as substrate. Samples were run in SDS-PAGE and stained with Coomassie to control for protein levels in each reaction and subsequently transferred to a PDVF membrane to assess H3K27 methylation levels by autoradiography. Results confirmed that the three mutant PRC2-EZH1 complexes have enhanced HMT activity relative to the PRC2- EZH1wt complex (Fig 3e-f). Interestingly, each variant reached this result by a slightly different mechanism. EZH1-p.Q731E was hyperactive regardless of the substrate (H3K27me0 or H3K27me2). EZH1-p.L735F was more efficient than EZH1wt methylating H3K27me0 but had similarly low activity on H3K27me2. Finally, as expected by the similarity with cancer associated EZH2-p.A677G mutations, EZH1-p.A678G was hyperactive on H3K27me2 but hypoactive on H3K27me0. This finding implies that EZH1-p.A678G leads to increased H3K27me3 only on nucleosomes harboring H3K27me2 deposited by an EZH1wt copy or by EZH2.

Collectively, these results demonstrate that missense EZH1 variants have a disruptive effect on EZH1 catalytic function. HMT assays further dissect the exact molecular mechanisms that create distinct EZH1 GOF variants. Combined with EZH1 LOF variants found in our recessive NDD cohort, our observations suggest that precise EZH1 activity regulation is critical for neural development and homeostasis.

### Precise EZH1 expression is necessary for maintaining the balance between neural progenitor proliferation and differentiation

Having established the molecular consequences of EZH1 variants, we sought to investigate how loss or gain of EZH1 impacts neural development. For this purpose, we took advantage of the chick embryo neural tube developmental model, which has been used traditionally to identify fundamental processes of neural development proven to be conserved in mammals. We first examined the expression of EZH1 in a 5 -day old chick embryo neural tube, which is comprised of neural progenitor cells in the ventricular zone (VZ) and differentiating neurons in the peripheric mantle zone (MZ). Immunofluorescence results revealed that EZH1 is mostly expressed in postmitotic neurons (Fig 4a).

**Figure 4:**
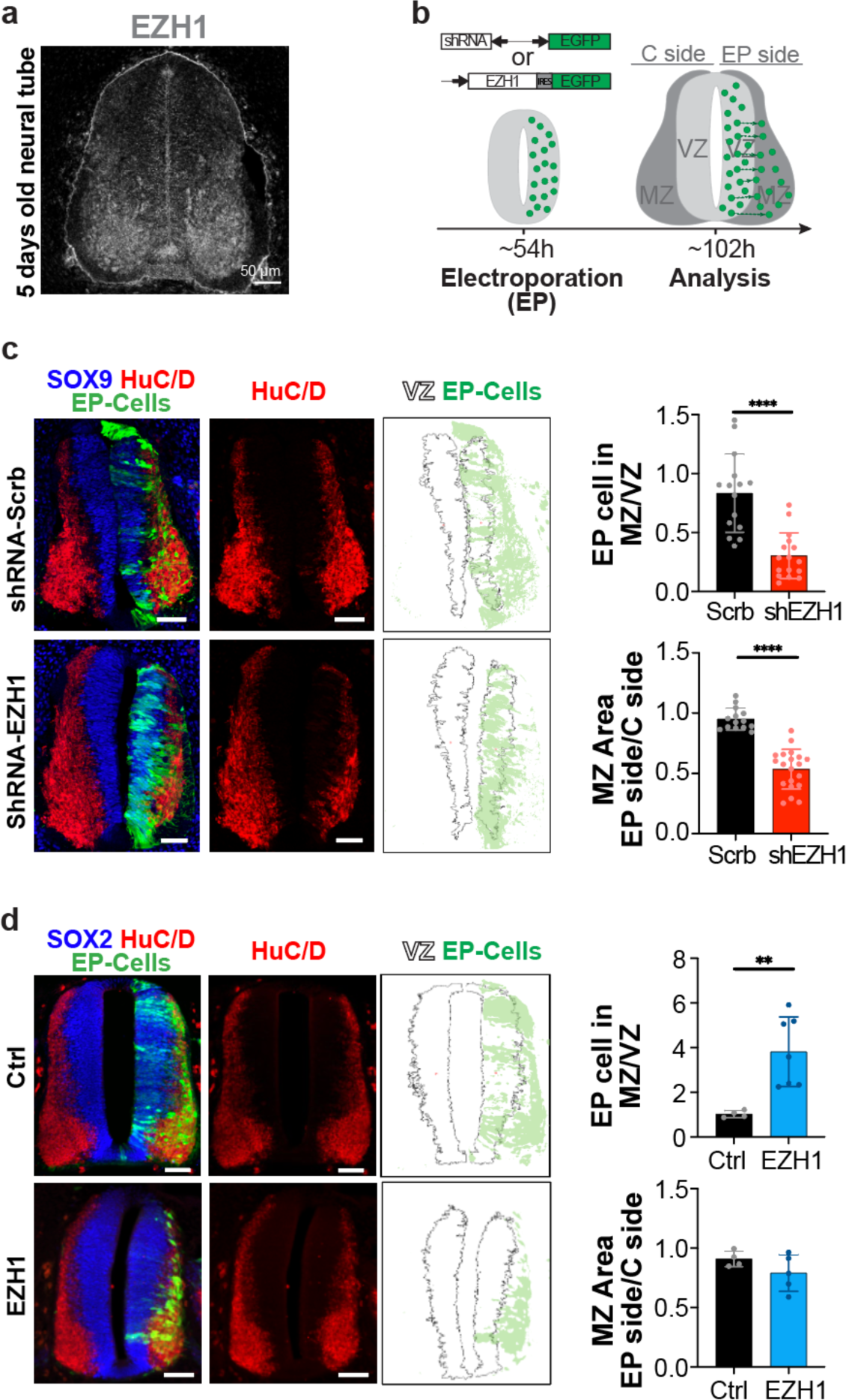
Gain and loss of EZH1 expression impairs neuronal differentiation during the chick embryo neural tube development. **a,** Representative image of embryonic day 5 chick embryo neural tube (NT) transversal sections immunostained with EZH1 antibodies showing predominant expression in post-mitotic neurons. **b,** Diagram summarizing the *in ovo* neural tube electroporation procedure. Electroporated (EP) cells are shown in green. VZ, ventricular zone (composed by neural progenitor cells). MZ, mantle zone (composed of differentiating neurons). **c,** Representative images of NT sections immunostained for SOX9 and HuC/D 48h after electroporation with DNA plasmids encoding EGFP and either a scramble shRNA or a mix of two EZH1 shRNAs. Top graph shows the ratio of EGFP positive EP cells located in the MZ vs in the VZ. Bottom graph shows the ratio of HuC/D stained MZ area between the electroporated and non-electroporated sides of the neural tubes. Data represents the mean +/-SD of n=14-21 sections from 2-4 embryos. **d,** Representative images of NT sections immunostained for SOX2 or HuC/D 48h after electroporation with DNA plasmids encoding EGFP or EZH1 and EGFP. Top graph shows the ratio of EP cells (EGFP^+^) located in the MZ vs in the VZ. Bottom graph shows the ratio of MZ stained area between the electroporated and non- electroporated sides of the NTs. Data represents the mean +/-SD of n=3-7 sections from 3-6 embryos . Significance was assessed by Student’s t-test for the ratio of MZ areas between EP and control side and by Mann-Whitney U test for MZ/VZ EP cell ratio. n.s.: p-value>0.05; *p-value<0.05, **p-value<0.01, ***p-value<0.001, ****p-value<0.0001.

Next, we analyzed the effect of EZH1 loss or gain during the chick embryo neural tube development. To attain EZH1 loss, we electroporated (EP) plasmids encoding shEZH1 and GFP into embryonic day 2 neural tubes and analyzed the tissue 48h later (Fig 4b-c). Compared to the scramble shRNA, the expression of two independent EZH1 shRNAs (shEZH1) successfully reduced EZH1 signal (Supplementary Fig 3). Furthermore, the proportion of shEZH1 EP cells (GFP+) migrating from the VZ (where they were when electroporated) to the MZ was half of that in controls. Consequently, shEZH1 electroporated neural tubes yielded less postmitotic neurons than the controls, as shown by the smaller HuCD stained area (Fig 4c). In contrast, the overexpression of EZH1 by electroporation of a plasmid also co-expressing GFP, increased 4- fold the proportion of EP cells localized in the mantle zone compared to controls (Fig 4d). Notably, mantle zone EP cells were HuCD positive, like the neighboring differentiating neurons. Although, the size of the MZ was not significantly altered in this case, this result shows that the overexpression of EZH1 induces the differentiation of neural progenitor cells. In aggregate, these data indicate that EZH1 is necessary to regulate the balance between neural progenitor cells proliferation and differentiation during neural development.

### EZH1 is expressed in developing and adult cerebral cortex

Genes associated with neurodevelopmental disorders are expressed in the human cerebral cortex, particularly during the critical developmental periods of neurogenesis and synaptogenesis^10, 43^. To ascertain if EZH1 expression fits with this expression pattern, we examined developing and adult human brain transcriptomic databases. RNA sequencing data from the Genotype-Tissue Expression (GTEx) portal showed that *EZH1* is similarly expressed across different regions of the adult human brain, including the cerebral cortex (Supplementary Fig 4a). We next explored the BrainSpan gene expression database^44^, which includes RNA sequencing data from the human cerebral cortex at several pre- and postnatal stages. The interrogation of *EZH1* retrieved a homogeneous constant expression across all the pre and postnatal stages. In contrast, *EZH2* showed a sharp drop during the developmental window that overlaps with neurogenesis, remaining low thereafter (Fig 5a). These data suggest a possible central role for EZH1 as the predominant H3K27 methyltransferase beginning at cortical neurogenesis stages, and therefore, a concomitant vulnerability of the developing cerebral cortex to EZH1 loss or gain of function.

**Figure 5:**
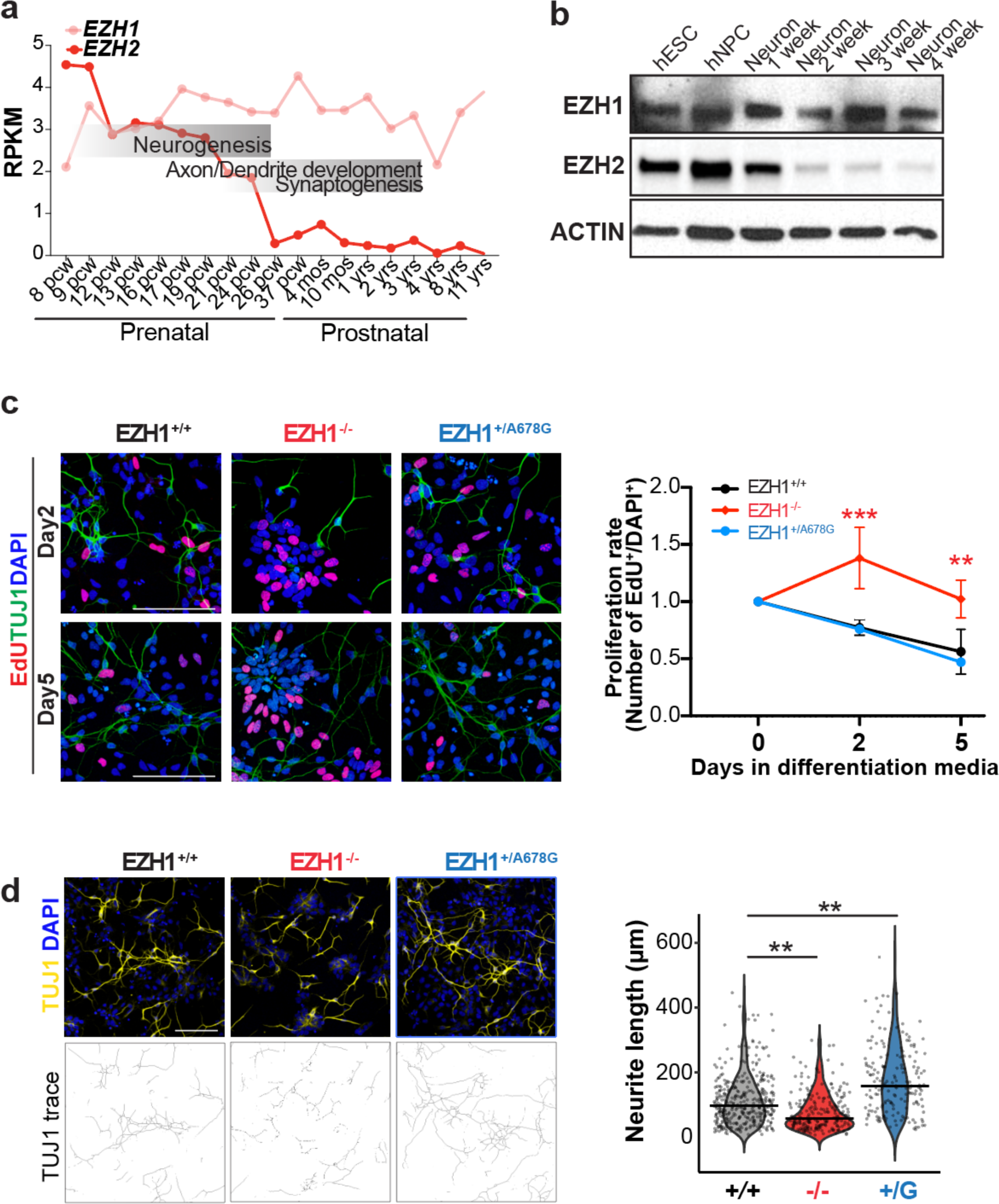
Loss and gain of function EZH1 variants, respectively delay and accelerate hPSC derived cortical neuron differentiation. **a,** *EZH1* and *EZH2* expression levels (RPKM) during pre- and postnatal cerebral cortex development. Data mined from mRNA-sequencing datasets in the Brainspan Atlas. **b,** Western blot analysis of EZH1 and EZH2 in hPSCs and derived cortical neural progenitors (hNPC) and neurons at the indicated week of differentiation. Actin is shown as loading control. **c,** Representative images of EdU labeling and TUJ 1 immunostainings upon 2 and 5 days differentiation of NPCs. Graphs show mean +/-SD of n=3 biological replicates. Statistical significance was assessed with two-way ANOVA with Tukey’s post hoc analysis test for multiple comparisons. **p-value<0.01***, p- value<0.001. **d,** Representative images of Tuj1 immunostaining upon 2 days differentiation of NPCs. Bottom panel shows traces of Tuj1 neurites. Violin plots show neurite length quantifications of 179-353 cells from 3 independent differentiations per genotype. One-way ANOVA with Dunnett’s post hoc analysis test for multiple comparisons. *p- value<0.05.

### EZH1 LOF and EZH1 GOF variants delay and promote cortical neuron differentiation respectively

Increasing evidence supports hPSC derived neurodevelopmental models as unique and versatile resources to study mechanisms of neurodevelopmental disorders^45–47^. Additionally, we verified that EZH1 and EZH2 expression dynamic is conserved in hPSC derived cortical neuron differentiation (Fig 5b). Thus, we generated isogenic hPSCs carrying a EZH1 LOF mutation (EZH1^-/-^) and one of the patients’ GOF variants (EZH1^+/A678G^) using CRIPSR/Cas9 genome engineering technology and subsequently validated editing, pluripotency and genome integrity (Supplementary Fig 4b-e). WB results confirmed loss of EZH1 in EZH1^-/-^ and intact EZH1 levels in EZH1^+/A678G^ (Supplementary Figure 4f).

To investigate the effects of EZH1^-/-^ and EZH1^+/A678G^ in cortical neuron differentiation, we first generated cortical neural progenitor cells (NPC) from hPSCs using standard protocols^48^. As expected, all NPCs expressed PAX6 and NESTIN (Supplementary Fig 4d). However, we noted that EZH1^-/-^ NPCs continue expanding even after differentiation induction, while EZH1^+/+^ and EZH1^+/A678G^ NPCs gradually stop proliferating. To validate this observation, we assessed the proliferation rates by EdU incorporation and labeling upon 0, 2 and 5 days of differentiation induction. Results confirmed that proliferation rates of EZH1^-/-^ NPCs are similar across the first 5 days differentiating, while EZH1^+/+^ and EZH1^+/A678G^ reduce proliferation rates after only 2 days in differentiation media (Fig 5c). Since this data suggests delayed differentiation of EZH1^-/-^ NPCs, we next assessed neurite length after tubulin III (TUJ1) immunostaining as a parameter of neuron maturation. Consistently, upon induction to differentiate, EZH1^-/-^ cells exhibited shorter neurites compared to wild type cells (Fig 5d). Interestingly, this analysis also revealed the opposite effect in EZH1^+/A678G^ cells, which showed longer neurites than wild type cells (Fig 5d). These data indicate that EZH1 LOF impairs neuronal differentiation while EZH1 GOF promotes maturation of differentiating neurons and suggest defects in neurogenesis timing.

### EZH1LOF and EZH1GOF variants alter the schedule of cortical projection neuron generation

While monolayer cortical neuron differentiations provide large amounts of NPCs and neurons ideal for robust mechanistic studies, they lack the spatiotemporal organization of the developing cerebral cortex. This deficiency hinders our ability to assess defects in neurogenesis regulation, such as asynchronies in timing and type of neuron generation. Instead, forebrain organoids recapitulate the temporal fate restriction coupled with laminar distribution of the developing cerebral cortex. In forebrain organoids, NPCs are arranged around a luminal structure in the ventricular zone (VZ) and they progressively give rise to deep layer neurons first and upper layer neurons, located at the periphery later^49^. Thus, to test if EZH1 mutations cause defects in cortical neurogenesis timing, we differentiated EZH1^-/-^ and EZH1^+/A678G^ hPSC using the sliced forebrain organoid protocol^49^. After a month in culture, all organoids showed neuroepithelial structures comprised by SOX2^+^ NPC containing VZs. While the thickness of the VZs was similar between EZH1^+/A678G^ and EZH1^+/+^ organoids, EZH1^-/-^ exhibited thicker VZs (Fig 6a). This data is consistent with prolonged proliferation rates we observed in EZH1^-/-^ NPCs monolayer cultures and supports deficits in neuronal differentiation.

**Figure 6:**
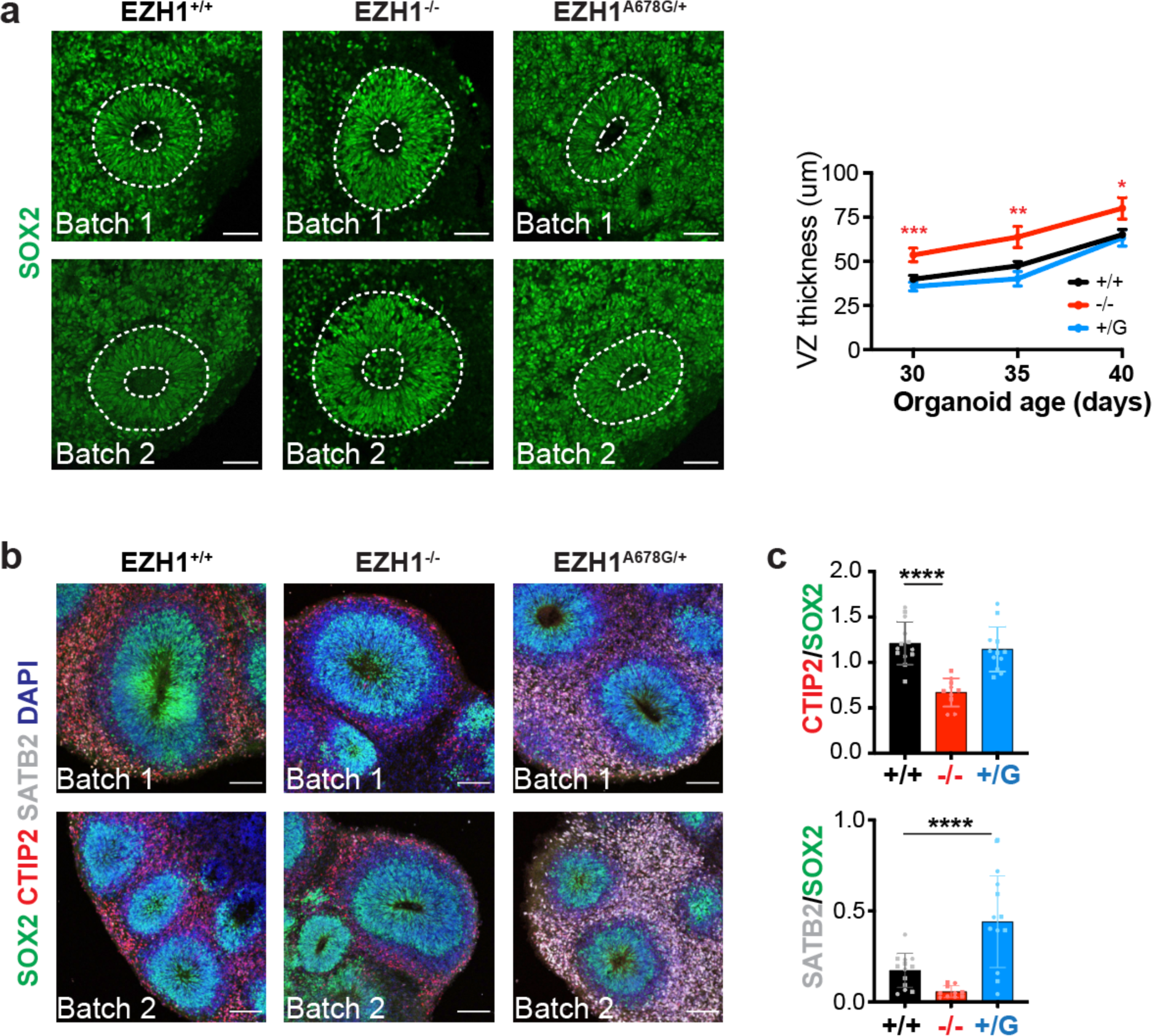
Cortical organoids derived from hPSCs carrying EZH1 LOF and GOF variants display abnormal timing of up and low layer cortical neuron generation. **a,** Representative images of 35-day old forebrain cortical organoids immunostained with SOX2. Graph shows mean +/-SEM of SOX2 immunostained VZ thickness in 30, 35 and 40-day old organoids (n=11-23 organoids per batch). **b,** Representative images of two batches of 60-day old forebrain cortical organoids immunostained with SOX2 (neural progenitor marker), CTIP2 (deep-layer cortical neuron marker), and SATB2 (upper-layer cortical neuron marker), showing less CTIP2 stained neurons in EZH1^-/-^ organoids and more SATB2 stained neurons in organoids carrying a gain of function variant (EZH1^+/A678G^). **c,** Graphs showing mean +/-SD of CTIP2/SOX2 (up) and SATB2/SOX2 (down) ratios in n=5-7 organoids per batch. Statistical significance was assessed using Nested ANOVA with Dunnett’s post hoc analysis test for multiple comparisons. ****p-value<0.0001.

To further test if EZH1 variants affect neuronal differentiation we next analyzed 60 day old forebrain organoids. At this age, wild type forebrain organoids were comprised by SOX2^+^ NPCs in the VZ, and CTIP2^+^ deep layer neurons around them. The first few SATB2+ upper layer neurons were also detected at this stage. Interestingly, the analysis of 60-day old EZH1^-/-^ and EZH1^+/A678G^ organoids revealed marked differences compared to wild type organoids. Specifically, EZH1^-/-^ organoids showed less CTIP2^+^ neurons (Fig 6b). In contrast, in EZH1^+/A678G^ organoids, the amounts of CITP2^+^ neurons were comparable to EZH1^+/+^ organoids, but they exhibited larger amounts of SATB2^+^ neurons (Fig 6b). To control for the intrinsic variability in the size of neuroepithelial structures between and within organoids we quantified the ratio of differentiated neurons (CITP2^+^ or SATB2^+^) over the SOX2^+^ NPCs in each neuroepithelial structure, which confirmed neurogenesis deficits in EZH1 LOF and GOF organoids (Fig 6c).

In summary, these results indicate that *EZH1* variants alter the timing of neurogenesis to distinct cortical neuron populations. Unscheduled neurogenesis events may ultimately result in defective development of cortical neuron networks causing the overlapping neurodevelopmental disorders associated with EZH1 LOF and GOF variants.

## Discussion

In this study, we uncover recessive and dominant *EZH1* variants as the cause of overlapping neurodevelopmental disorders and demonstrate an evolutionarily conserved function of EZH1 regulating neuronal differentiation. The recessive variants are all LOF, including two homozygous nonsense mutations predicted to impair EZH1 expression in 7 children from 3 independent consanguineous families. We also identified a sporadic case with biallelic splice and deletion variants confirmed to cause loss of EZH1. These findings are consistent with a homozygous frameshift *EZH1* variant (*c.237delT*; p.L80Sfs*6) recently identified in two siblings through an independent genetic study of recessive intellectual disabilities^50^. Like patients in our cohort, the two siblings in this study were diagnosed with intellectual disability and showed dysmorphic faces. The recurrence of *EZH1* LOF variants in unrelated individuals with similar phenotypes independently identified, further supports the association of EZH1 in the etiology of a previously undefined NDD.

The remaining 9 patients in our cohort all carry heterozygous missense *EZH1* variants likely occurred *de novo*. To our knowledge, missense *EZH1* variants have never been associated with a human disease before, except for somatic EZH1 p.Q571R and p.Y642F mutations that are recurrent in minimally invasive thyroid and parathyroid tumors^51–53^. Although it is unclear how these EZH1 mutations contribute to tumor formation or progression, the molecular characterization of EZH1 p.Q571R variant suggests a GOF effect^52^ reminiscent of the missense mutations in our NDD cohort. GOF mutations in EZH2 are also common in human cancers that motivated the development of EZH inhibitors as therapeutic agents^41^. Notably, one such mutation (i.e. EZH2 p.A677G) is equivalent to EZH1 p.A678G variant found in our patient P4^40, 54, 55^. Although none of our patients show malignancies, the molecular coincidence corroborates the GOF nature of the missense EZH1 mutations in our NDD cohort. Nonetheless, it is possible that each missense variant affects EZH1 function in different ways. In line with this idea, our *in vitro* HMT assays revealed distinct mechanisms for H3K27 hypermethylation in three of the EZH1 missense variants that alter conserved SET domain residues (Fig 3). Furthermore, two missense variants fall far from the catalytic SET domain and near the EZH1 MCSS-SANT2L loop. Remarkably, the MCSS-SANT2L loop has recently been associated with an EZH1 specific ability to bind and condense chromatin in a methyltransferase activity independent manner^38^. This finding suggests a potential effect of variants p.R406H and p.E435D on EZH1 mediated chromatin condensation that warrants further investigation.

Likewise, further work is required to better understand how EZH1 LOF and GOF converge on overlapping neurodevelopmental phenotypes. Counterintuitively, our work shows apparent opposing effects of EZH1 LOF and GOF in neural development. While EZH1 LOF delays neuronal differentiation, EZH1 GOF promotes it. Nonetheless, these opposing defects are consistent with an asynchronous neurogenesis that is emerging as a shared neurodevelopmental defect causing genetically heterogeneous NDDs^46^. Like in KMT5B, CHD8 and ARID1B happloinsufficient brain organoids reported in these studies, EZH1 LOF delays cortical projection neuron neurogenesis. Although the effect of EZH1 GOF is opposite and different in type of neurons affected, it similarly causes defects in the schedule of neurogenesis that could lead to abnormal neuronal network activity and explain the overlapping neurodevelopmental phenotypes observed in EZH1 LOF and GOF patients. Regardless, our work demonstrates that cortical neurogenesis is vulnerable to EZH1 dosage and suggests that EZH1 LOF and GOF may converge altering the epigenetic transcriptional regulation of neurodevelopmental programs in a temporal and lineage specific manner.

Although other core PRC2 subunits have been genetically implicated in developmental syndromes^56^ there are marked differences between these syndromes and the EZH1 associated NDDs identified here. Specifically, the hallmark of patients with pathogenic variants in *EZH2*, *EED* and *SUZ12* is overgrowth with ID, while most patients in our EZH1 cohort have short or normotypic statures. Additionally, despite the overlap in ID manifestations, the neurobiological mechanisms causing neurodevelopmental defects are likely different. While pathogenic mechanisms that cause ID in PRC2 associated overgrowth syndromes remain unclear, it is well established that PRC2-EZH2 maintains proliferative neural progenitor pools and dynamically regulates neural cell fate transitions during neurogenesis^57–59^. In contrast, our work shows that EZH1 promotes neuronal differentiation and regulates the timing of neurogenesis of cortical projection neurons. This is consistent with a previously reported function of EZH1 in synaptic spine maturation^33^ and with the switch from EZH2 to EZH1 predominance that occurs as neural progenitors differentiate (Fig 5a). Thus, together, these data support that EZH1 and EZH2 have non-redundant functions in neural development.

NDDs caused by chromatin dysregulation are attractive candidates for treatment with so-called epigenetic drugs. Recent years have witnessed an exponential increase in the development of epigenetic drugs, mostly purposed for the treatment of human cancers. Although these drugs are not recognized as treatment options for NDDs, the similarity of EZH1 GOF variants with those found in EZH2 associated cancers suggest a potential route for epigenetic recovery for EZH1 GOF using EZH1/2 inhibitors. Furthermore, H3K27me3 demethylase inhibitors may have the potential to counterbalance EZH1 or PRC2 LOF variants in NDDs. Treating NDDs presents several challenges, including the prenatal onset of the disease and the requirement for brain penetrant compounds. However, there are successful examples of postnatal reversal of neurodevelopmental phenotypes in animal models of Rett, Kabuki and Angelman syndrome^60–62^, raising hope for therapeutic targeting of PRC2 NDDs. Overall, our work uncovers a critical role of EZH1 in human neural development, provides a molecular diagnosis for patients with previously undefined NDDs and offers experimental models to test the therapeutic potential of epigenetic drugs to treat phenotypes of NDDs.

## Methods

### Identification of pathogenic EZH1 variants

Affected individuals were identified through diagnostic clinical practice or connections through GeneMatcher^30^ and Deciphering Developmental Disorders Research Study^31^ repositories. Genomic DNA was extracted from peripheral blood samples of the probands and parents and siblings when available. Exome or genome sequencing was performed with a variety of standard capture kits and libraries sequenced with available sequencing platforms. Data analysis was performed to assess quality of sequence reads and variants filtered as with several pipelines previously described^63–65^. Variants were reported according to standardized nomenclature recommended by the Human Genome Variation Society^66^ on EZH1 transcript GenBank:

NM_001991.5. Genomic sequencing data in gnomAD was used to determine the frequency of identified variants in the population and missense tolerance score retrieved from Metadome^67^. Conservation was determined by alignment of EZH1 and EZH2 protein sequences from different species using ClustalW.

### Patient consent

Human subject studies were approved by the local IRBs, including the Children’s Hospital of Philadelphia, Boston Children’s Hospital, University College London, Guy’s Hospital, Kennedy Krieger Institute, King Faisal Specialist Hospital & Research Center and University of Alabama in Birmingham. Informed consent for participation, phenotyping, sample collection and publication of images was obtained through the referring clinical teams.

### Structural characterization of the missense variants

For each variant we built a three-dimensional model using the package MODELLER1. The structure of EZH1 (PDB: 7KSO) was used as a template for the E438D variant. For variants R406H, R728G, Q731E, L735F, because 7KSO did not cover their locations, we followed a simple, two-step protocol. First, we created a model of the native EZH1, using the experimental structure of EZH2 (PDB: 5HYN) as a template. Then, we utilized the resulting model to build the structure of the desired variants. Variant A678G was also analyzed using this model, because it includes an H3K27me3 mimetic peptide which allows us to assess the effect of A678G on the catalytic pocket. In the case of L735F, to illustrate the potential stacking interaction between the aromatic side chain of the phenylalanine amino acid and the SAH molecule, we added local information from the structure of the SMYD3-SAM complex (PDB: 3MEK).

### RNA extraction, cDNA generation and PCR

Total RNA was extracted from patient and control LCLs using TRIzol Reagent (Invitrogen # 15596026) and purified using RNeasy Plus Mini Kit (Qiagen). For splice site analysis, directed reverse transcription was performed using 0.5 μg of total RNA and 0.2 µM of reverse *EZH1* or *GAPDH* primers (Supplementary Table 2) with SuperScript™ III First-Strand Synthesis System (Invitrogen # 18080051). Standard PCR protocol was performed using 0.5 µM forward and reverse primers (Supplementary Table 3). Amplicons were ran in a 2% agarose gel and imaged using BioRad Gel Doc EZ Imager. For quantification of transcripts by RT-qPCRs, 0.5-2 μg of total RNA was retrotranscribed with oligodTs and SuperScript™ III First-Strand Synthesis System following standard protocol. Next, qPCR was performed in a 10 µL reaction volume containing 4.6 µL of cDNA sample, 0.5 µM of forward and revers primers (Supplementary Table 4) and 1X Power SYBR Green PCR Master Mix in a Biorad CFX384 Touch Real-Time PCR. Primers were designed using Benchling’s Molecular Suite Primer3 program or NCBI Blast.

### In vitro histone methyltransferase assay

Standard HMT assays were performed in a total volume of 15 mL containing HMT buffer (50mM Tris-HCl, pH 8.5, 5 mM MgCl2, and 4mM DTT) with 10 mM of ^3^H-labeled S-Adenosylmethionine (1:20 of ^3^H-labeled:unlabeled), 15 mM of H3K27me3, indicated amount of recombinant human PRC2-EZH1 complexes and 120 nM of either unmodified or H3K27me2 mononucleosomes. The reaction mixture was incubated for 60 min at 30°C and stopped by the addition of 4 mL SDS buffer (0.2 M Tris-HCl, pH 6.8, 20% glycerol, 10% SDS, 10 mM b-mercaptoethanol, and 0.05% Bromophenol blue). After HMT reactions, samples were incubated for 5 min at 95°C and separated on SDS-PAGE gels. The gels were then subjected to Coomassie blue staining for protein visualization or wet transfer of proteins to 0.45 mm PVDF membranes (Millipore). The radioactive signals were detected by exposure on autoradiography films (Denville Scientific).

### ReNcell VM Culture

ReNcell VM human neural precursor cells were plated onto Corning® Matrigel® hESC-Qualified Matrix -coated T75 cell culture flasks (CELLSTAR® Filter Cap Cell Culture Flasks) and cultured in ReNCells media containing 1:1 ratio of N2 and B27 media supplemented with 20 ng/ml Fibroblast Growth Factor (bFGF) and 20 ng/ml Epidermal Growth Factor (bEGF). N2 media contains DMEM/F12 (Gibco #11330032), 1X N-2 neural supplement (Gibco #17502048), 5 μg/ml Insulin (MilliPore Sigma #I9278), 1X GlutaMAX™ (Gibco # 35050061), 100 μM MEM Non-Essential Amino Acids Solution (Gibco #11140050), 100 μM β-Mercaptoethanol (Gibco #21985023), 1 mM Sodium Piruvate (Gibco #11360070) and 1% Penicillin-Streptomycin (Gibco #15140122). B27 media contains Gibco Neurobasal™ medium (Gibco #21103049) supplemented with 1X B-27 neural supplement (Gibco #17504044), 1X GlutaMAX™and and 1% Penicillin-Streptomycin. Cell culture media was changed every 2 days and cells passaged using accutase (Gibco #A1110501) when reached 90% confluency.

### Lentiviral Production and Transduction of ReNCells

Full-length cDNAs of human wildtype or mutant EZH1 were cloned between the EF1A promoter and T2A-EGFP sequence in pLVs vector (VectorBuilder). For lentiviral production, HEK293-T cells were plated at 1x10^6^ cell/well density in a well of a 6-well plate with 2 ml of media (1X DMEM (Gibco #11995073), 10%FBS (Tissue Culture Biologicals #101), and 1%Penn/Strep (Gibco #15140122)). The following day, cells were transfected using polyethyleneimine (PEI) with 1.06 μg MDLg/RRE (gag/pol expression plasmid), 0.57 μg MD2.G (VSV-G envelope expressing plasmid), 0.4 μg pRSV-Rev (rev expression plasmid), and 1.6 μg of transfer vector (pLVs) per well. 48 h later 2 mL of media with lentiviruses were collected and filtered through 0.45 μm pore filters. 100-200 μL of filtered lentiviral media was added to ReNcells seeded the day before at 800,000 cells/well density in a well of a 6 well plate. ReNcells were incubated with lentiviruses for 48 h after which time media was replaced with fresh ReNcell media and cells cultured for 5 days before sorting with BD FACS Aria Fusion flow cytometer.

### Chick embryo neural tube electroporation

Eggs from white Leghorn chickens were incubated at 38.5 °C and 70% humidity. Embryos were staged according to the method of Hamburguer and Hamilton (1951). *In ovo* electroporations were performed at stage HH14 (∼54h of incubation with 22 somites) with purified plasmid DNA at 3 μg/ul in H2O and 50 ng/ml Fast Green. EZH1 overexpression plasmids were obtained from Vector Builder and EZH1 shRNAs expressed by cloning them into the pSHIN plasmid^68^. Electroporations were performed delivering five pulses of 50 ms 20-25 V. Embryos were collected 48h post electroporation.

### Pluripotent stem cell culturing and differentiation

H9 human pluripotent stem cells (hPSCs) (WiCell WA09) and edited lines were cultured on Matrigel (Corning, 354277) coated plates in mTeSR1 media (STEMCELL Technologies, # 85850). 10 μM Rho-associated protein kinase (ROCK) inhibitor (Y-27632 Tocris, Cat. No. 1254) was only added into the culture media overnight when thawing the ESC/IPSCs. The medium was changed every day and cells were passaged when they reached 70% confluence, approximately every 5- 7 days, using Versene (Gibco, 15040066) at a 1:15 or ReleSR (STEMCELL Technologies, #100- 0484) at a 1:30 ratio.

### Generation of genetically modified hPSCs by CRISPR-CAS9

The 20bp sgRNA sequences were cloned into the PX458 vector (Addgene, #48138). Single stranded DNA oligonucleotides (ssODNs) were synthesized by Integrated DNA Technologies (IDT). ROCK inhibitor was added to the culture media 24h prior to electroporation. On the day of electroporation, Human Stem Cell Nucleofector Kit 1 (LONZA, #VPH-5012) was prepared based on the manufacturer’s protocol. ESC/IPSCs were singularized with Accutase (Gibco, A1110501) and counted with Countess II Automated Cell Counter. To generate heterozygous clones, 3.5 μg PX458 plasmid (Addgene Plasmid #48138, from Feng Zhang) containing target sgRNA sequence and 1 μg ssODN containing missense variants and two synonymous variants that disrupt PAM sequence plus 1 μg of an ssODN containing only the two synonymous variants were diluted in 100 μl transfection solution, and mixed with 1.2x10^6^ singularized cells. To generate KO clones, 3.5 μg plasmid was diluted in 100 μl transfection solution, and mixed with 1.2x10^6^ singularized cells and Amaxa Human Stem Cell Nucleofector Kit I. Each mixture was transferred to one nucleofection cuvette. B-016 nucleofector program was selected on Amaxa Nucleofector and the current was applied to the cell/DNA suspension cuvette. After electroporation, the cells were transferred to a Matrigel coated well with 10 μM ROCK inhibitor in the media. 24h after electroporation, cells were fed with fresh mTeSR1 and 1% Penicillin-Streptomycin (P/S) (Gibco, 15140122). 48h after electroporation, the cells were singularized with Accutase, resuspended in mTeSR1 with 1%P/S, and filtered through 35μm nylon mesh cell strainer cap tube (Falcon 352235). 1,000 – 2,000 GFP positive cells were sorted through the BD FACSAria™ Fusion Flow Cytometer, collected in mTeSR1 with CloneR supplement (STEMCELL Technologies, #05888), and seeded on a 10cm dish coated with Matrigel. Single colonies were manually picked on day 8 to 10 and half of the colony was transferred to Matrigel coated 96-well plates and the other half processed for genomic DNA extraction followed by PCR and Sanger sequencing. Clones successfully edited were identified and two of them selected for experiments. The primers, sgRNAs, and ssODNs used for genome editing are listed in Supplementary Table 5.

### Cortical neuron differentiation

Cortical neurons were generated as previously described with few modifications^48^. Briefly, hPSC colonies were disassociated with accutase, and seeded at ∼50,000 cells/cm^2^ density on Matrigel coated wells in mTeSR1 + 5µM ROCK inhibitor. Next day media was replaced by neural induction media (50% DMEM/F12 and 50% Neurobasal, supplemented with N2 (Gibco, 17502048, 1:100), B27 (Gibco, A3582801, 1:50), GlutaMAX (Gibco, 35050061, 1:100), and 50µM BME (Gibco, 21985023) 100nM LDN193189 (STEMCELL Technologies, #72147) and 10µM SB431542 (Stemgent, 04-0010-10)). 5 days later cells were singularized with accutase and plated at a ∼250,000 cells/cm^2^ density in Matrigel coated wells, in neural induction media for additional 6 days. Next day cells were singularized again and plated at a ∼250,000 cells/cm^2^ on matrigel coated wells in neural progenitor expansion media (50% DMEM/F12 and 50% Neurobasal, supplemented with 1xN2, 1xB27, 1xGlutaMAX, 50µM BME and 20ng/ml bFGF). Media was replaced everyday. On Day 25, NPCs were singularized and plated on 100µg/ml Poly-L-ornithine (Sigma-Aldrich, P3655) + Matrigel coated wells in neuronal differentiation media (50% DMEM/F12 and 50% Neurobasal supplemented with 1xB27, 1xGlutaMAX, 50µM BME, 10ng/ml BDNF (Biothchne, 248-BD/CF), 10ng/ml GDNF (Biothchne, 212-GD/CF), and 1µg/ml laminin (Gibco, 23017015). Thereafter, ½ media was replaced with fresh media every other day.

### Sliced cortical organoids differentiation

Forebrain organoids were generated as previously described^49^. On Day 0, iPSC colonies were detached by ReLeSR and aggregated to form EBs by Aggrewell (STEMCELL Technologies, Catalog # 34811). The following day, EBs were resuspended and transferred to 6-well plate (Corning Costar) rotating at 110 rpm, containing DMEM/F12, 20% KnockOut Serum Replacement, 1X Non-essential Amino Acids, 1X GlutaMax, 1 μM LDN193189, 5 μM SB-431542 and 2 μg/mL heparin (STEMCELL Technologies). On Day 6, half of the medium was replaced with induction medium consisting of DMEM/F12, 1X N2 Supplement, 1X Non-essential Amino Acids, 1X GlutaMax, 1 μM CHIR99021 and 1 μM SB-431542. On Day 7, organoids were embedded in Matrigel and cultured stationarily in Low-attachment plate (Corning Costar) in the induction medium. On Day 14, organoids were mechanically dissociated from Matrigel by manual pipetting in a 10 mL pipette tip. Organoids were transferred to 6-well plate rotating at 110 rpm, containing differentiation medium consisting of DMEM:F12, 1X N2 and B27 Supplements, 1X 2- Mercaptoenthanol, 1X Non-essential Amino Acids, and 2.5 mg/ml human Insulin. From Day 35 to Day 70, 1% Matrigel was supplemented in differentiation medium. At Day 70, differentiation medium was exchanged with maturation medium consisting of Neurobasal medium, 1X B27 Supplement, 1X 2-Mercaptoenthanol, 0.2 mM Ascorbic Acid, 20 ng/ml BDNF, 20 ng/ml GDNF, and 1 μM Dibutyryl-cAMP.

### Fluorescent Activating Cell Sorting (FACS)

Cells were disassociated with Accutase™ (Gibco # A1110501) and pelleted at 200 g for 5 minutes. Pellet was resuspended with 1 mL of 1X PBS with 10% FBS and filtered through a 35 µm nylon mesh cell strainer cap into a 5mL Falcon® Round-Bottom Tubes (Corning #352235). Cells were loaded into the BD FACS Aria Fusion flow cytometer coupled with a BD FACSDiva Software. After excluding doublets or non-viable cells, GFP+ events were collected into a tube containing 0.5ml of cell specific media.

### Chromatin Immunoprecipitation, Sequencing, and Analysis

Cells were fixed with 1% formaldehyde for 10 min at room temperature. Fixation was quenched with glycine (0.125 M). Briefly, cross-linked chromatin was fragmented by sonication carried out using a Bioruptor to generate an average fragment size of 200-500 bp. Chromatin was purified by centrifugation for 30 min, at 20 000g and 4°C. Purified chromatin was further diluted in immunoprecipitation (IP) buffer (0.01% SDS, 1.1% Triton X-100; 1.2 mM EDTA pH 8.0; 16.7 mM Tris–HCl pH 8.1; 167 mM NaCl) and incubated overnight with 1 μg of H3K27me3 antibody (Cell Signaling (D5A7)). Protein A-bound beads were added and immunocomplexes were washed once with buffers TSE I (0.1% SDS; 1% Triton-X100; 2 mM EDTA pH 8.0; 20 mM Tris–HCl pH 8.1; 150 mM NaCl), TSE II (0.1% SDS; 1% Triton-X100; 2 mM EDTA pH 8.0; 20 mM Tris–HCl pH 8.1; 500 mM NaCl) and TSE III (0.25 M LiCl; 1% NP-40; 1% sodium deoxycholate; 1 mM EDTA pH 8.0; 10 mM Tris–HCl pH 8.1) and twice with TE buffer (10 mM Tris–HCl pH 8.1 and 1 mM EDTA). De-crosslinking was carried out for 4 hrs at 65°C in elution buffer (1% SDS, 0.1 M NaHCO3). DNA fragments were purified using the QIAquick PCR Purification procedure to remove fragments >10kb. DNA libraries were constructed with the TAKARA ThruPLEX® DNA-Seq Kit and the Illumina-compatible TAKARA DNA Single Index Kit. DNA libraries were quantified using a high sensitivity Chip on Bioanalyzer (Agilent) and sequenced at 40 million 150 bp pair-end reads per replicate in the NovaSeq 6000 (Illumina).

ChIP-seq reads were aligned to the hg19 reference genome using bowtie2 with parameters: -q --local --no-mixed --no-unal –dovetail^69^. Uniquely aligned reads (mapping quality score >= 20) and concordant alignments were kept for downstream analysis using command: samtools -q 20 -f 0x2. H3K27me3 peaks were called using MACS2 with parameters: --broad --keep-dup all -p 1e-5 -- broad-cutoff 1e-5^70^. ChIP-seq signal was normalized to sequencing depth using deepTools: bamCoverage --normalizeUsing CPM^71^. ChIP-seq signal around peaks was computed and visualized using deepTools computeMatrix and plotHeatmap functions, respectively.

### Western Blotting

For histone-specific Western Blotting, histones were enriched by lysing cells in an acid lysis buffer (10nM HEPES, 1.5 mM MgCl2, 10mM KCl). For all other Western Blots protein lysates were generated using RIPA Buffer (Cell Signaling #9806). Protein lysates and histone extractions were separated by SDS-PAGE, transferred to a PVDF membrane, and detected by Western blotting. The membrane was blocked with 5%BSA for 1 hr at room temperature, incubated with one of the following antibodies (EZH2 [Cell Signaling (D2C9)], 1:1,000; H3K27me3 [Cell Signaling (D5A7)], 1:1,000; β-actin: [GenScript (A00702)], 1:5,000; H4: [Abcam(ab10158] 1:10,000) in 5%BSA at 4C overnight. The next day the membrane was washed three times with 7 mL of 1X TBS with 0.01% Tween20, incubated for 1 hr with the corresponding secondary antibodies (Invitrogen anti-Rabbit HRP, anti-Rabbit [# 31458] and anti-Mouse HRP [# SA1-100]), and washed three times with 1X TBS with 0.01% Tween20. The membranes were developed using Pierce™ ECL Western Blotting Substrate kit (Thermo Scientific #32106). ImageJ software was used to quantify each protein band and normalize it to a loading control (β-actin or H4).

### Immunofluorescence staining

For chick embryo neural tube analyses, embryos were fixed for 2h at 4°C in 4% paraformaldehyde and immunostaining was performed in 50 µm free floating sections cut with a vibratome. Sections were permeabilized with PBS-0.1% Triton X-100, blocked with BSA solution and immunostained with the following primary antibodies: mouse anti-HuC/D (Millipore A-21271) or mouse anti-NeuN (Chemicon MAB377) for MZ staining and rabbit anti-SOX2 (Abcam AB97959) or rabbit anti-SOX9 (kindly provided by James Briscoe) to label the VZ. Alexa Fluor 555, and Cy5 conjugated secondary antibodies were obtained from Invitrogen. Sections were stained with 1µg/ul DAPI and mounted in Mowiol (Sigma-Aldrich). Images were acquired with the Zeiss Lsm780 confocal system. The effects of EZH1 shRNA electroporation were assessed by measuring the area occupied, respectively, by the VZ (formed by SOX2+ or SOX9+ progenitors) or the MZ (formed by HuC/D+ or NeuN neurons) in chick neural tube transversal sections. The analysis was adapted from^60^. Briefly, VZ or MZ channels were split and measured using the ImageJ software. After producing z-stack maximal projection images, a threshold was applied to define the areas in each side of the neural tube. VZ or MZ areas were measured in both the control and the electroporated side of the neural tubes by a particle analysis using a pixel^2 size ranging from 1000 to infinity. The data are presented as ratios of the area ± SD obtained by standardizing the values of the electroporated (EP) side with the corresponding values of the non-EP side of the neural tube. Similarly, the ratio of cells in the VZ and MZ was produced by manually counting the number of GFP+ cells in the VZ and MZ in each neural tube.

hPSCs and derived neural cells were plated onto Matrigel coated glass coverslips (Electron Microscopy Sciences 72290-04). Cells were fixed with 4% PFA for 10 min at room temperature, permeabilized and blocked in blocking buffer (5% goat serum (Sigma-Aldrich, G9023) and 0.1% Triton X-100 in PBS) for 1 h at room temperature, and immunostained over night at 4 °C with the following antibodies: 1:500 SSEA (Abcam ab16287), 1:500 OCT4 (Abcam ab19857), 1:300 PAX6 (Biolegend 901302), 1:2000 Nestin (Millipore Sigma MAB326), 1:1000 TUJ1 (Abcam ab18207). Next day, cells were incubated for 1h at room temperature with 1:500 of Alexa Fluor 488, 555 or 633 – conjugated secondary antibodies and 0.1 μg/ml Hoechst 33342 and mounted with Mowiol mounting medium (sigma #81381). For EdU (5-ethynyl-2’-deoxyuridine, Invitrogen E10187) labeling, cells were treated with 10 μM EdU for 30 min. After fixation for 10 min at RT with in 4% PFA, EdU labeling was performed using Click-it plus EdU cell proliferation kit following the manufacturer’s instructions (Invitrogen C10638). Antibody co-staining was performed after EdU staining as indicated above. Images were acquired with Leica SP8 confocal microscope and analyzed in FIJI ImageJ. TUJ1 trace images were generated by FIJI ImageJ Process, Binary, skeletonize.

Whole organoids were fixed in 4% formaldehyde in Phosphate Buffered Saline (PBS) for 30 mins at room temperature. Organoids were washed with PBS and then immersed in 30% sucrose solution overnight. Organoids were embedded in tissue freezing medium and sectioned with a cryostat (Leica) at 30 μm thickness. For immunostaining, cryosectioned slides were washed with PBS. Tissues were permeabilized with 0.5% Triton-X in PBS for 1 hr and blocked with blocking medium consisting of 10% donkey serum in PBS with 0.05% Triton-X (PBST) for 30 mins. Primary antibodies (SOX2 (R&D Systerms, AF2018), CTIP2 (Abcam, ab18465), SATB2 (Abcam, ab51502)) diluted in blocking solution were applied to the sections overnight at 4 °C. After washing with PBST for 3 times, secondary antibodies (AlexaFluor 488, 546, or 647 -conjugated donkey antibodies (Invitrogen)) diluted in blocking solution were applied to the sections for 1.5 hr at room temperature. DAPI (Invitrogen) was added for 5 mins (Invitrogen) at the end of incubation. Finally, sections were washed with PBST for 3 times before mounting. Images were captured by a confocal microscope (Zeiss LSM 800). Sample images were prepared in ImageJ (NIH). Total CTIP2+ or SATB2+ cells were counted in each image and normalized to SOX2+ cell numbers. Analyses of ventricular zone thickness was performed as previously described^49^.

## Data Availability

The data generated to support the findings of the study are included in the manuscript or available from the corresponding authors upon reasonable request.

## Acknowledgments

We thank the Center for Cellular and Molecular Therapeutics Stem cell core and Center for Applied Genomics sequencing cores at Children’s Hospital of Philadelphia (CHOP) for their support with stem cell methods and sequencing technologies. The authors extend their appreciation to the invaluable contribution of Hessa Alsaif, other clinical coordinators, patients and their families to this study. This work was supported by the CHOP/UPENN IDDRC-New Program Development award, CHOP-Junior Faculty Pilot Program award, Margaret Q Landerbergen Foundation Award and NIH/NINDS 1R01NS119699-01A1 (to N.A.), BFU2015- 69248-P and PGC2018-096082-B-I00 from the Spanish Ministry of Economy to (M.A.M.B.), Alabama Genomic Health Initiative F170303004 through University of Alabama at Birmingham (to A.C.H., M.T.), PID2019-111217RB-I00 Spanish Ministerio de Ciencia e Innovación (to X.D.L.C.), the King Salman Center For Disability Research Group no RG-2022-010 (to F.S.A.), ID2019-110157RA-I00 (to M.S.), FPU Spanish Ministry of Education and Science predoctoral fellowship (to R.F.) and 2021 FISDU 00400 (to P.E-B.). This research was made possible through access to the data and findings generated by the 100KGP and the DDD. The 100KGP is managed by Genomics England Limited (a wholly owned company of the Department of Health and Social Care). The 100KGP is funded by the National Institute for Health Research and NHS England. The Wellcome Trust, Cancer Research UK and the Medical Research Council have also funded research infrastructure. The 100KGP uses data provided by patients and collected by the National Health Service as part of their care and support. The DDD study presents independent research commissioned by the Health Innovation Challenge Fund [grant number HICF-1009-003]. This study makes use of DECIPHER (http://www.deciphergenomics.org), which is funded by Wellcome.

## Disclosures

Erin Torti is an employee of GeneDX.

---

**Supplementary Figure 1:**
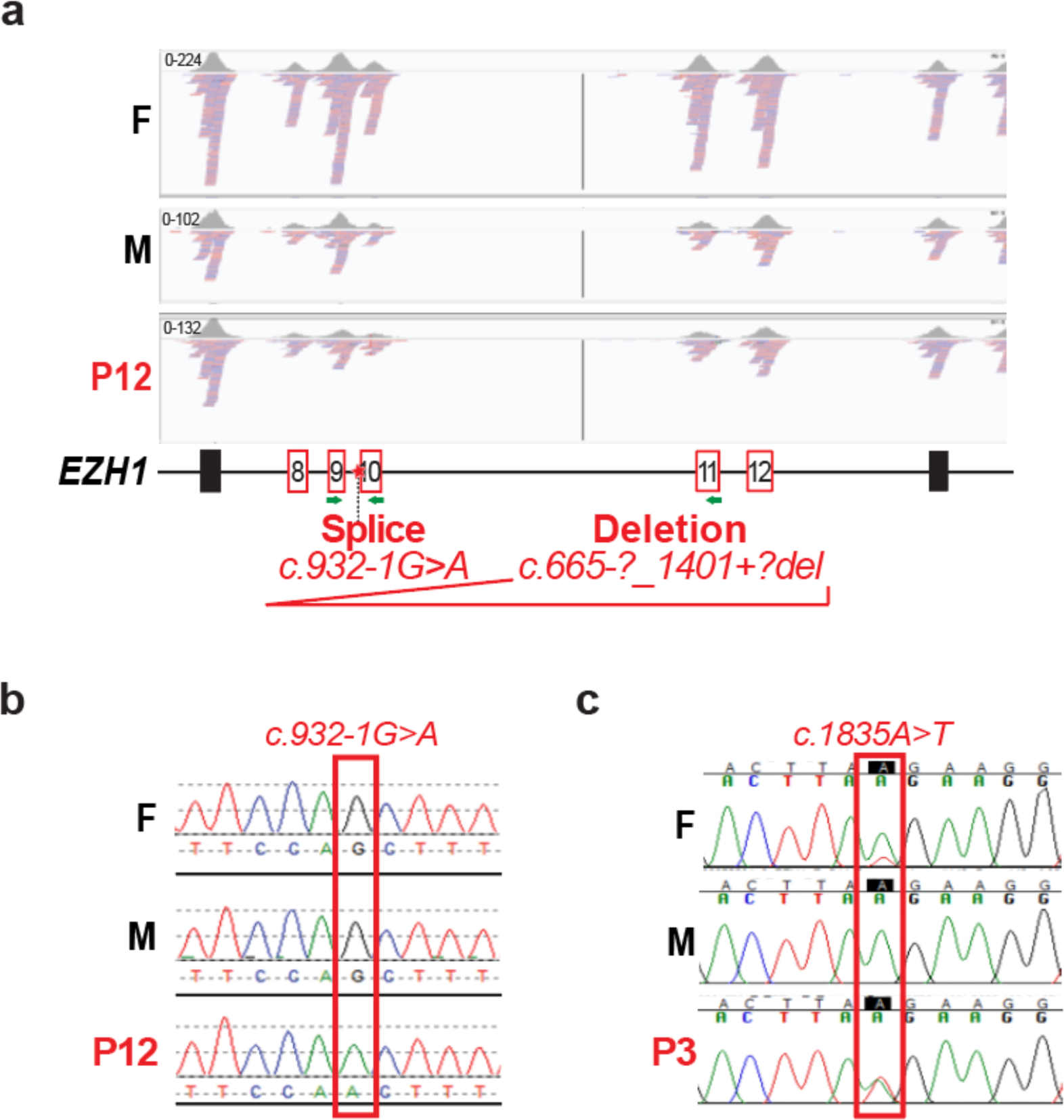
Exome sequencing and Sanger sequencing of two affected families carrying EZH1 variants and additional photos of affected individuals. **a,** IGV screenshot depicting exon sequencing reads corresponding to *EZH1* exon 7-13 in the parent F, parent M and the only affected individual (P12) of a non-consanguineous family. The parent M and the affected child (P12) show half number of reads covering exon 8-12, which suggests a monoallelic deletion of this region in the parent M that was transmitted to the affected child. **b,** Sanger sequencing of the parent F, M, and P12 showing the splice variant only in the affected child, thus suggesting a *de novo* origin. Note that the variant falls within the region deleted in the alternate allele. **c,** Sanger sequencing of DNA extracted from the parent F, M and affected child (P3) showing that the pathogenic variant is in heterozygosity in P3 and detected at low frequency in the parent F that is likely mosaic for the variant. **d,** Face and body photos of patients with EZH1 variants at various ages.

**Supplementary Figure 2:**
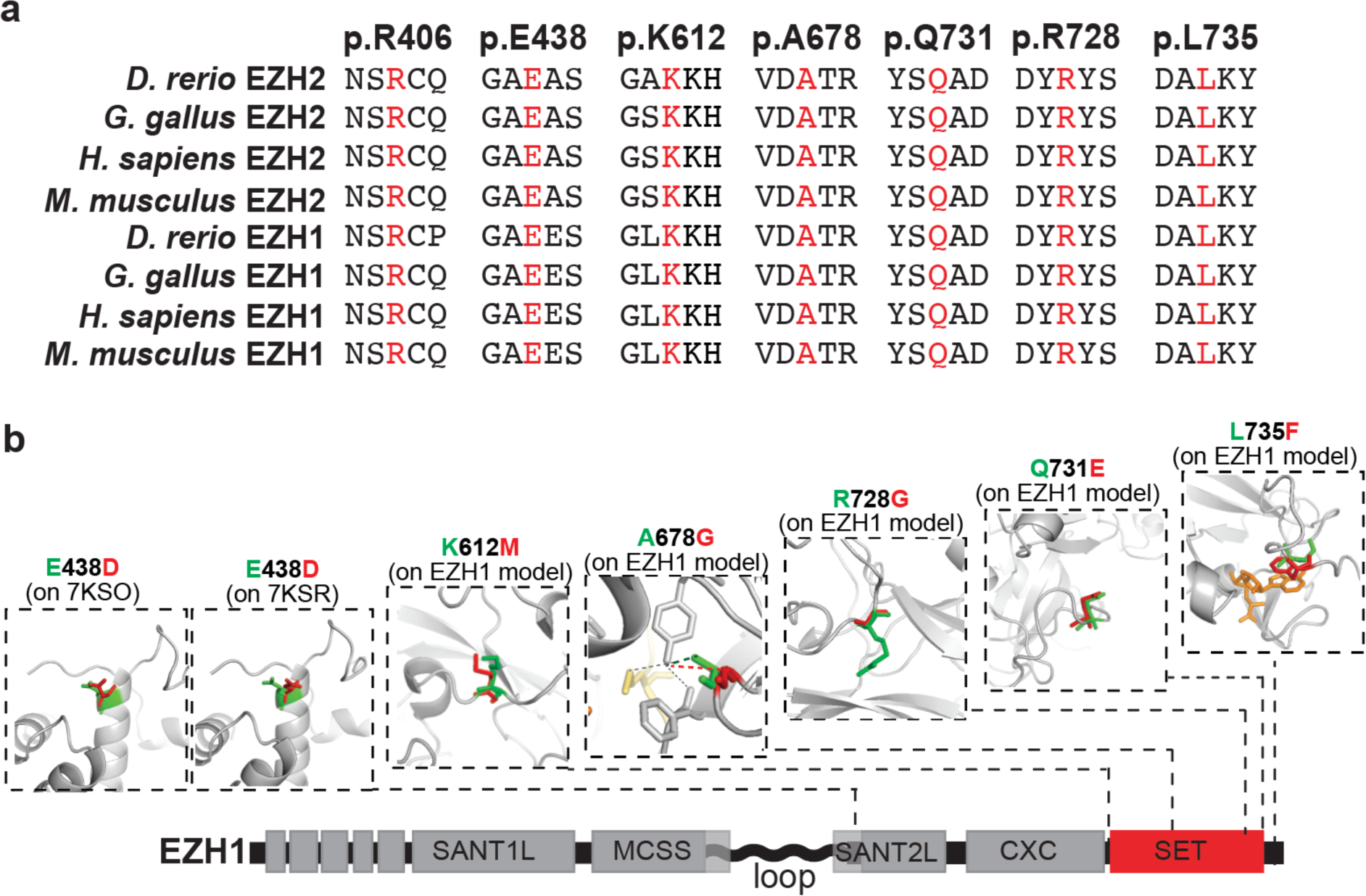
Missense EZH1 variants affect conserved residues and likely impact the interaction of EZH1 with H3K27, SAM or nucleosomes. **a,** Alignment of human EZH1 and EZH2 protein sequences and their orthologs show that missense variants identified in affected individuals in this study affect residues conserved from zebrafish to humans. **b,** Molecular modeling of missense EZH1 variants (red) overlapped with the native residue (green) on the experimental 3D structure of EZH1 bound (7KSO) and unbound (7KSR) to nucleosomes for p.E428D, or on the computationally predicted EZH1 structure modeled using the experimental structure of EZH2 (PDB: 5HYN) as a template for K612M, A678G, R728G, Q731E, and L735F variants. The yellow structure in A678G image represents the K27 in the histone tail. The orange structure in the L735F image represent SAM, the methyl donor molecule.

**Supplementary Figure 3:**
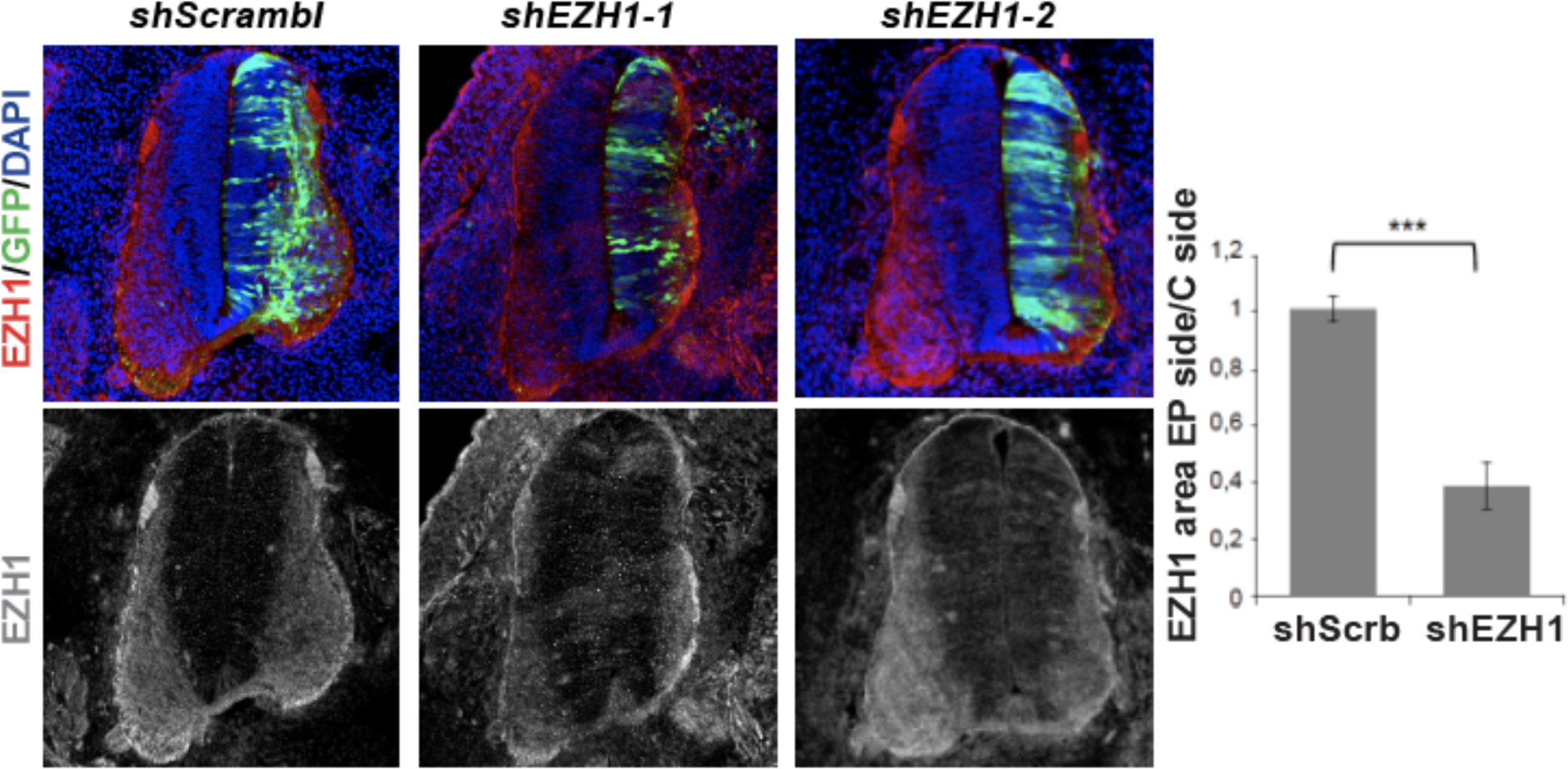
*in ovo* electroporation of EZH1 shRNA. Representative images of EZH1 immunostained neural tubes previously electroporated with a DNA plasmid encoding EGFP and a scramble shRNA or two different EZH1 shRNAs. Graph depicts the ratio of EZH1 stained area between the electroporated (EP side) and non- electroporated sides (C side) of the neural tubes from n=4-6 sections of 3 embryos. Student’s t-test. ***p-value<0.001.

**Supplementary Figure 4:**
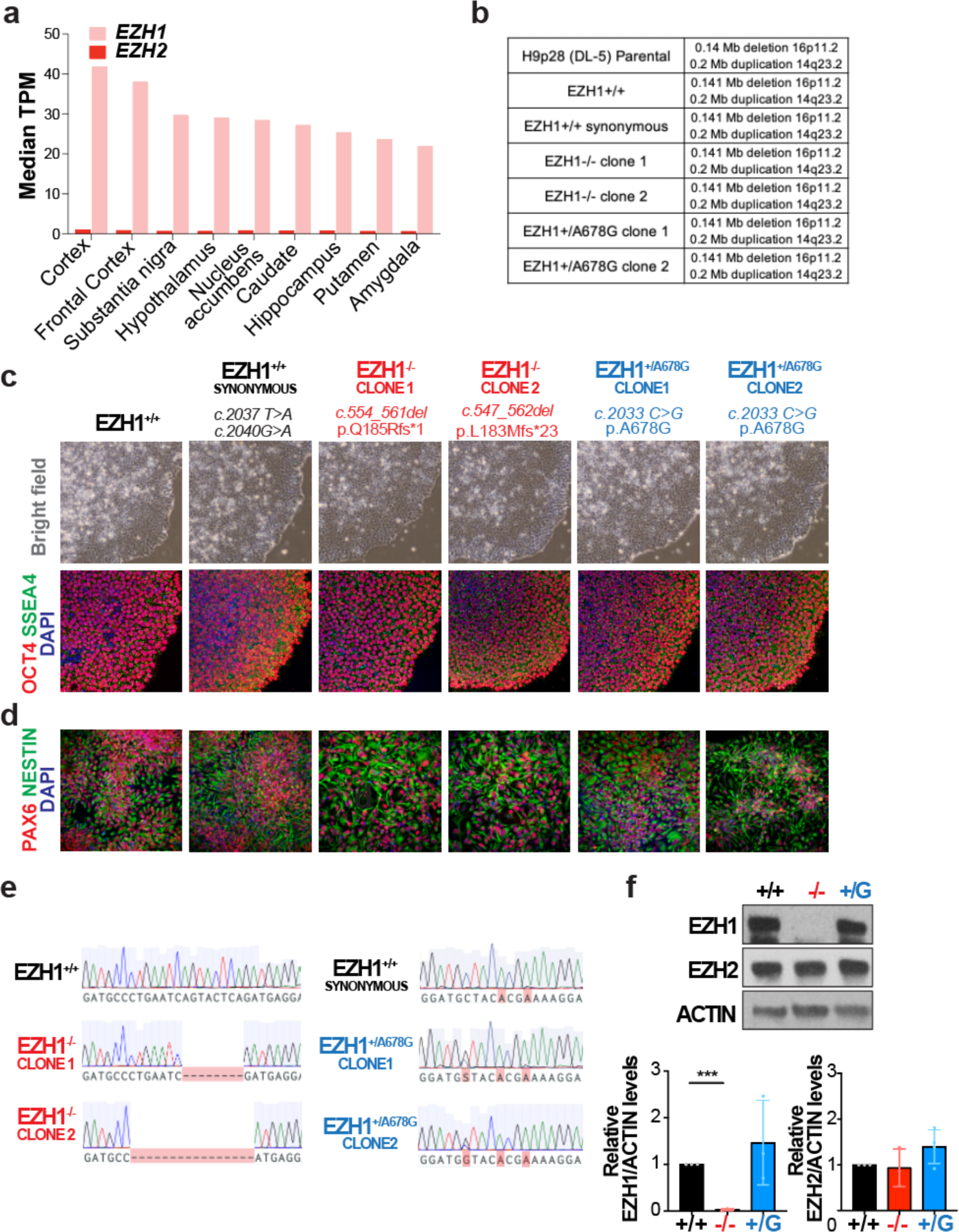
Human brain EZH1 expression and validation of iPSC editing. **a,** *EZH1* and *EZH2* expression levels (median TPM) on the indicated regions of the adult human brains. Data mined from mRNA- sequencing datasets in GTEX. **b,** Results from CNV analysis of the parental H9 hPSCs, non-edited H9 clone, H9 edited to carry EZH1 synonymous variants, two clones carrying EZH1 loss of function variant (EZH1^-/-^) or one of the patients’ GOF variants (EZH1^+/A678G^). **c,** Representative bright field images and immunostainings for pluripotency markers OCT4 and SSEA4 of hPSCs carrying EZH1 loss or gain of function variants introduced by CRIPSR/Cas9 genome editing. **d,** Representative images of hPSC-derived neural progenitor cells immunostainings with PAX6 and NESTIN. **e,** Sanger sequencing chromatograms showing *EZH1* variants in edited hPCS clones. **f,** Western blot analysis of EZH1 and EZH2 in hPSCs showing undetectable levels of EZH1 in hPSCs carrying a homozygous loss of function variant (EZH1^-/-^) and intact levels in hPSCs carrying a gain of function variant (EZH1^+/A678G^). EZH2 levels are similar in all. Actin is shown as loading control. Graphs show mean +/-SD of n=3 independent replicates. Statistical significance was assessed with one-way ANOVA with Dunnett’s post hoc analysis test for multiple comparisons. ***p-value<0.001.

**Supplementary Table 1:**
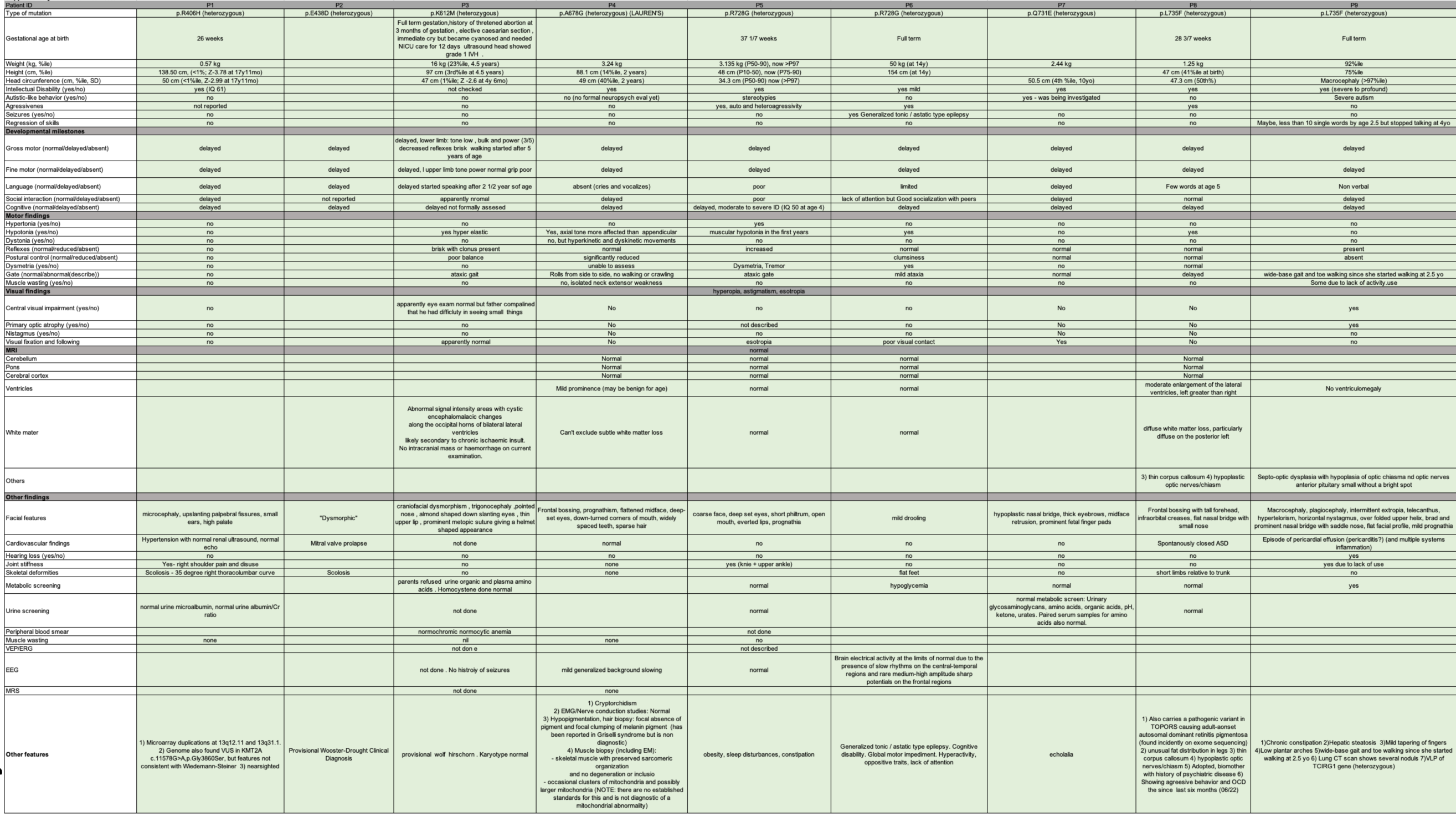
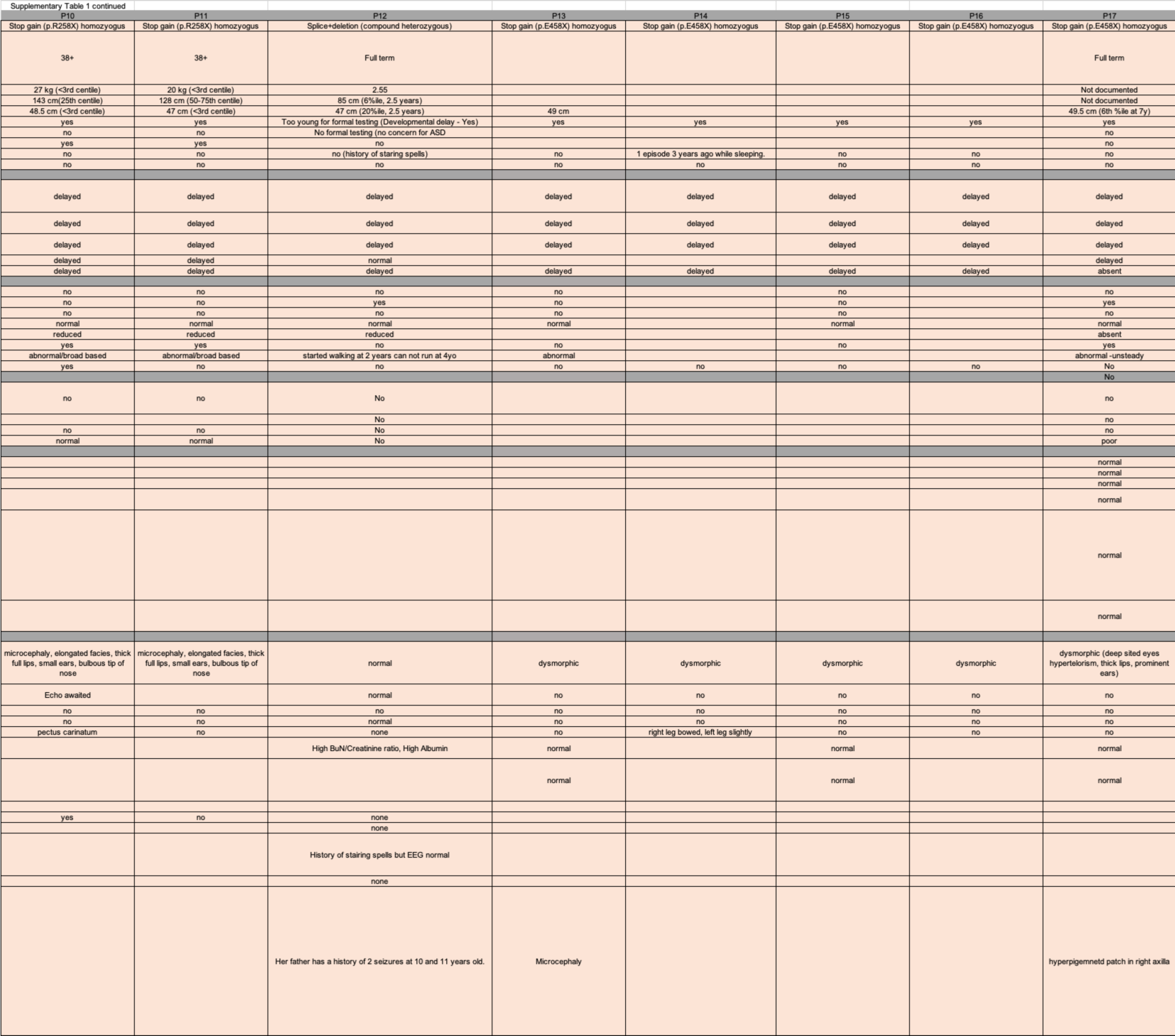
Clinical features

**Supplementary Table 2:**
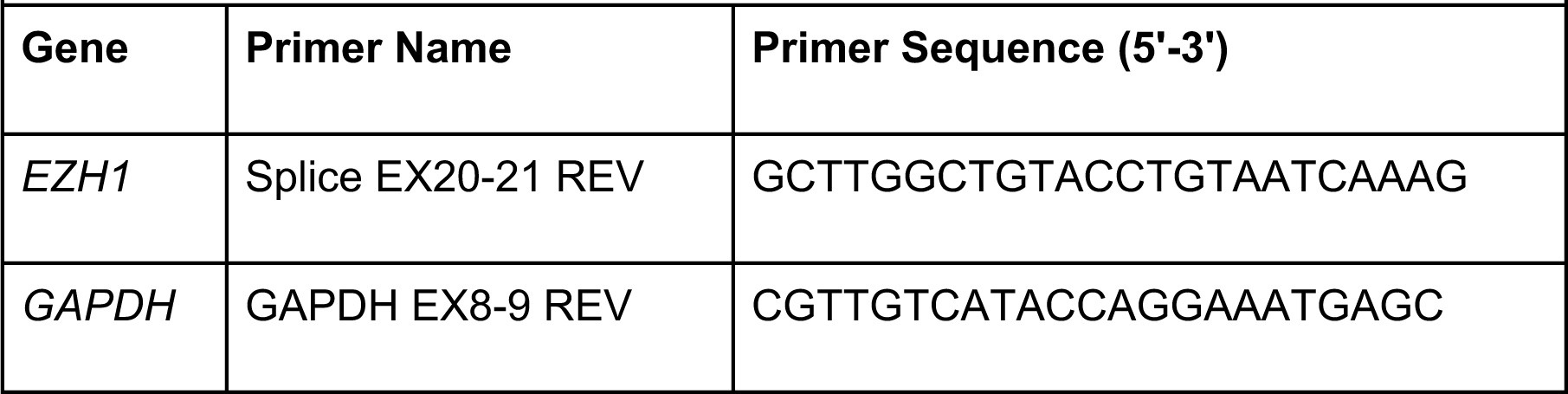
Primers to generate *EZH1/GAPDH* cDNA / RT-PCR

**Supplementary Table 3:**
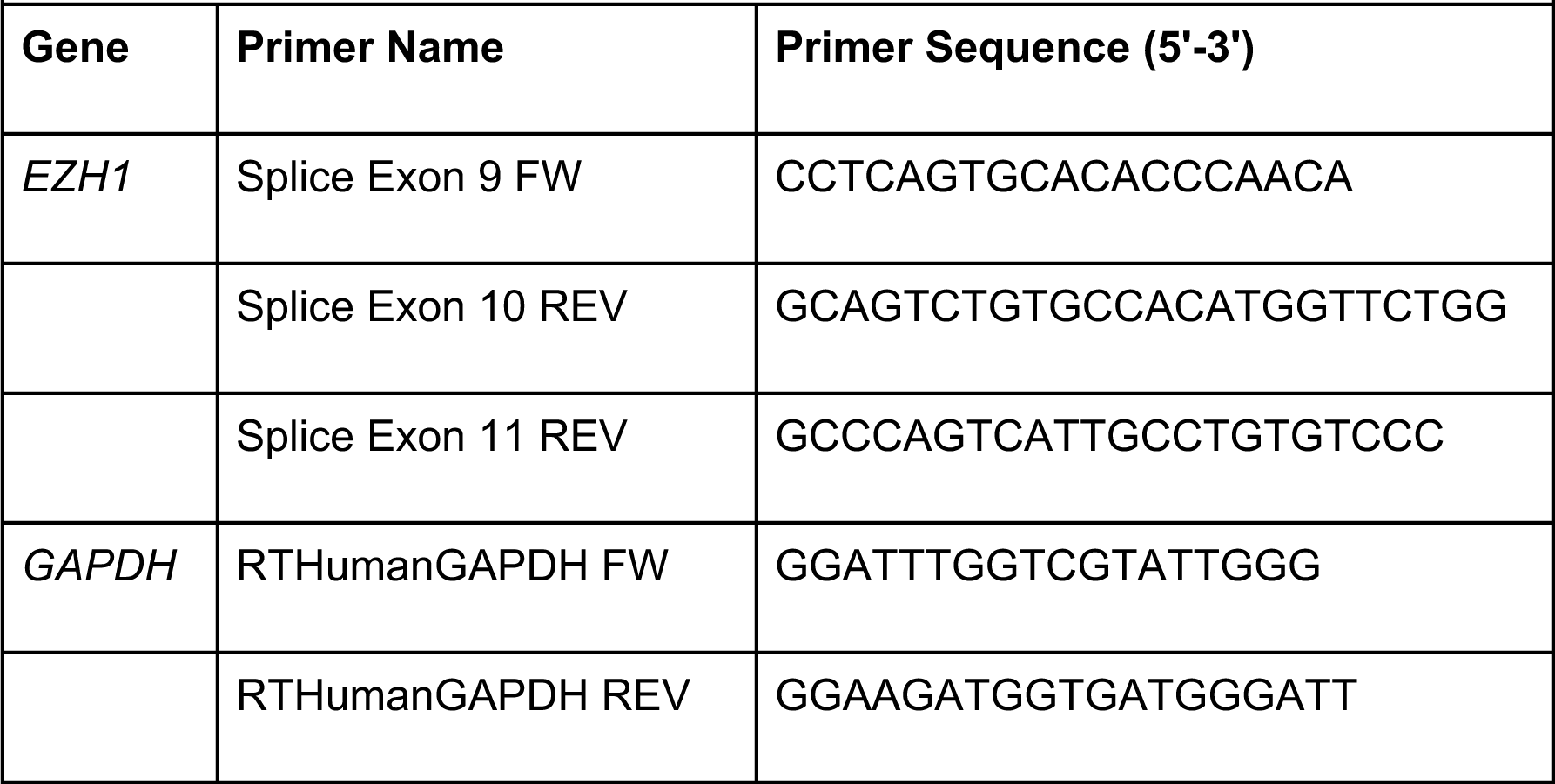
PCR Primers targeting Splice (P12) Mutation Site

**Supplementary Table 4:**
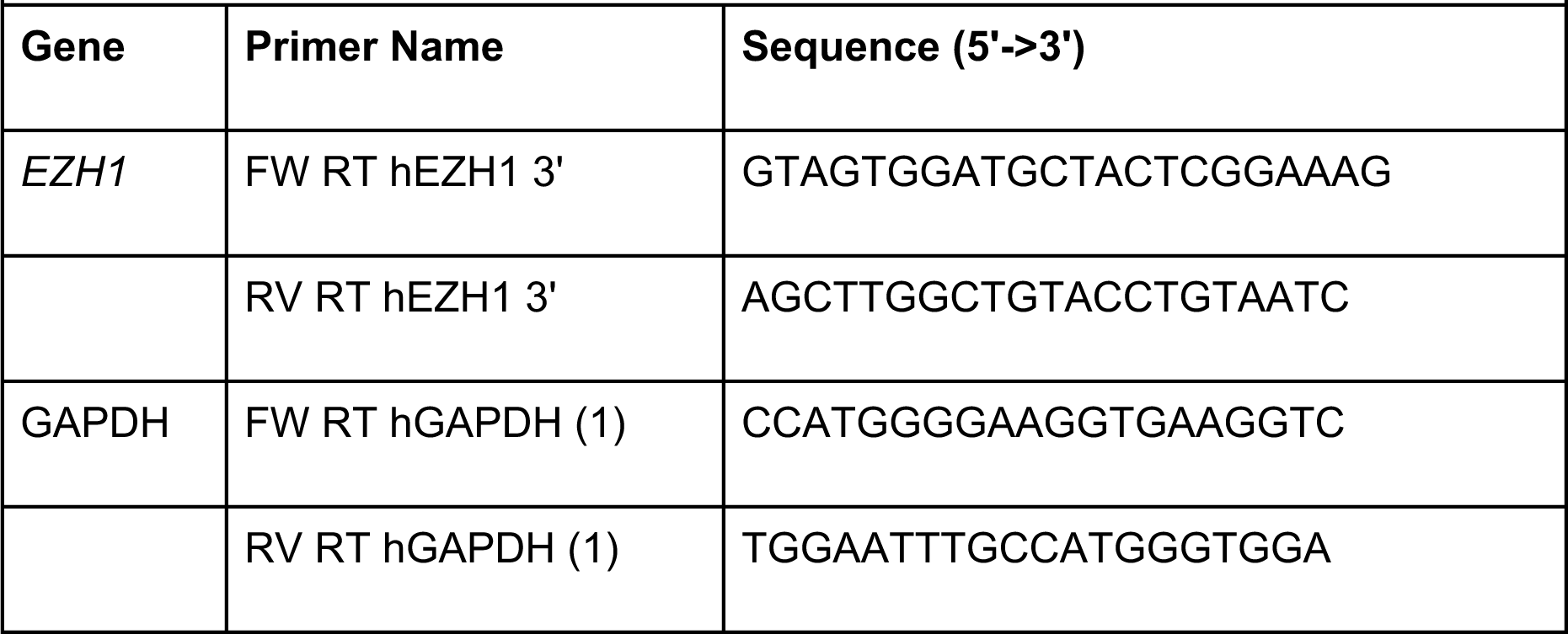
Primers used for RT-qPCR on Splice (P12) Samples

**Supplementary Table 5:**
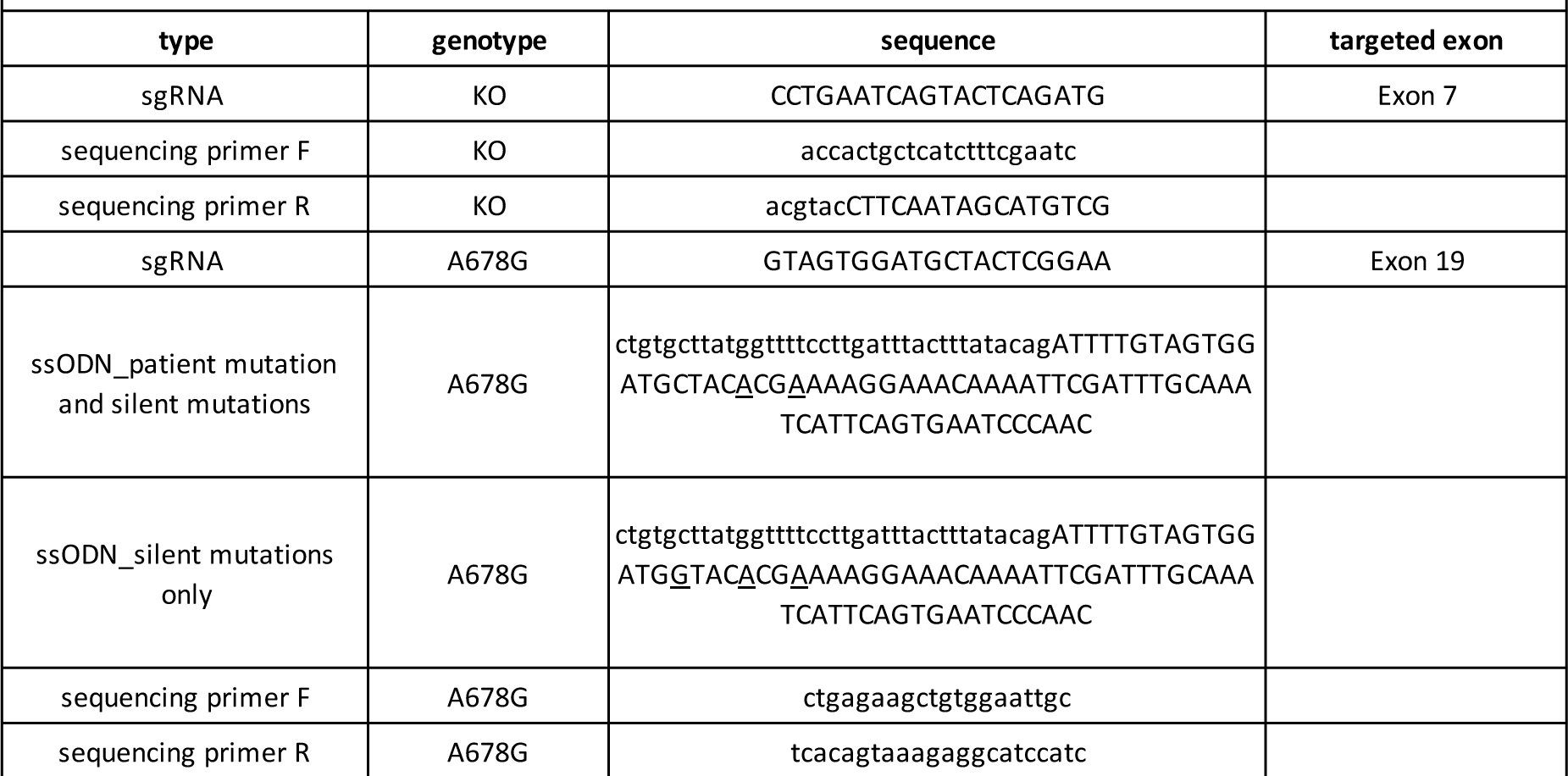
Oligonucleotides used for hPSC editing and testing

## Notes

### Competing Interest Statement

Erin Torti is an employee of LLC: GeneDx, LLC

### Author Declarations

All the human subject studies included here were approved by their local IRBs, including the Childrens Hospital of Philadelphia, Boston Childrens Hospital, University College London, Guys Hospital, Kennedy Krieger Institute, King Faisal Specialist Hospital & Research Center and University of Alabama in Birmingham. Informed consent for participation, phenotyping, sample collection and publication of images was obtained through the referring clinical teams.

## References

1. Wright, C. F. et al. Genetic diagnosis of developmental disorders in the DDD study: a scalable analysis of genome-wide research data. Lancet 385, 1305–1314, doi:10.1016/S0140-6736(14)61705-0 (2015).

2. Wilfert, A. B., Sulovari, A., Turner, T. N., Coe, B. P. & Eichler, E. E. Recurrent de novo mutations in neurodevelopmental disorders: properties and clinical implications. Genome Med 9, 101, doi:10.1186/s13073-017-0498-x (2017).

3. Wang, T. et al. Large-scale targeted sequencing identifies risk genes for neurodevelopmental disorders. Nat Commun 11, 4932, doi:10.1038/s41467-020-18723-y (2020).

4. Deciphering Developmental Disorders, S. Prevalence and architecture of denovo mutations in developmental disorders. Nature 542, 433–438, doi:10.1038/nature21062 (2017).

5. Kaufmann, P., Pariser, A. R. & Austin, C. From scientific discovery to treatments for rare diseases - the view from the National Center for Advancing Translational Sciences - Office of Rare Diseases Research. Orphanet J Rare Dis 13, 196, doi:10.1186/s13023-018-0936-x (2018).

6. Ciptasari, U. & van Bokhoven, H. The phenomenal epigenome in neurodevelopmental disorders. Hum Mol Genet 29, R42–R50, doi:10.1093/hmg/ddaa175 (2020).

7. Deevy, O. & Bracken, A. P. PRC2 functions in development and congenital disorders. Development 146, doi:10.1242/dev.181354 (2019).

8. Tatton-Brown, K. et al. Mutations in Epigenetic Regulation Genes Are a Major Cause of Overgrowth with Intellectual Disability. Am J Hum Genet 100, 725–736, doi:10.1016/j.ajhg.2017.03.010 (2017).

9. De Rubeis, S. et al. Synaptic, transcriptional and chromatin genes disrupted in autism. Nature 515, 209–215, doi:10.1038/nature13772 (2014).

10. Satterstrom, F. K. et al. Large-Scale Exome Sequencing Study Implicates Both Developmental and Functional Changes in the Neurobiology of Autism. Cell 180, 568–584 e523, doi:10.1016/j.cell.2019.12.036 (2020).

11. Yu, J. R., Lee, C. H., Oksuz, O., Stafford, J. M. & Reinberg, D. PRC2 is high maintenance. Genes Dev 33, 903–935, doi:10.1101/gad.325050.119 (2019).

12. Xiao, Z. G. et al. The Roles of Histone Demethylase UTX and JMJD3 (KDM6B) in Cancers: Current Progress and Future Perspectives. Curr Med Chem 23, 3687–3696, doi:10.2174/0929867323666160725093522 (2016).

13. Eich, M. L., Athar, M., Ferguson, J. E., 3rd & Varambally, S. EZH2-Targeted Therapies in Cancer: Hype or a Reality. Cancer Res 80, 5449–5458, doi:10.1158/0008-5472.CAN-20-2147 (2020).

14. Cyrus, S., Burkardt, D., Weaver, D. D. & Gibson, W. T. PRC2-complex related dysfunction in overgrowth syndromes: A review of EZH2, EED, and SUZ12 and their syndromic phenotypes. Am J Med Genet C Semin Med Genet 181, 519–531, doi:10.1002/ajmg.c.31754 (2019).

15. Conway, E., Healy, E. & Bracken, A. P. PRC2 mediated H3K27 methylations in cellular identity and cancer. Curr Opin Cell Biol 37, 42–48, doi:10.1016/j.ceb.2015.10.003 (2015).

16. Arnaud, M. et al. [Kabuki syndrome: Update and review]. Arch Pediatr 22, 653–660, doi:10.1016/j.arcped.2015.03.020 (2015).

17. Miyake, N. et al. MLL2 and KDM6A mutations in patients with Kabuki syndrome. Am J Med Genet A **161A**, 2234–2243, doi:10.1002/ajmg.a.36072 (2013).

18. Stolerman, E. S. et al. Genetic variants in the KDM6B gene are associated with neurodevelopmental delays and dysmorphic features. Am J Med Genet A 179, 1276–1286, doi:10.1002/ajmg.a.61173 (2019).

19. Lee, C. H. et al. Allosteric Activation Dictates PRC2 Activity Independent of Its Recruitment to Chromatin. Mol Cell 70, 422–434 e426, doi:10.1016/j.molcel.2018.03.020 (2018).

20. Margueron, R. et al. Ezh1 and Ezh2 maintain repressive chromatin through different mechanisms. Mol Cell 32, 503–518, doi:10.1016/j.molcel.2008.11.004 (2008).

21. Shen, X. et al. EZH1 mediates methylation on histone H3 lysine 27 and complements EZH2 in maintaining stem cell identity and executing pluripotency. Mol Cell 32, 491–502, doi:10.1016/j.molcel.2008.10.016 (2008).

22. Lee, C. H. et al. Distinct Stimulatory Mechanisms Regulate the Catalytic Activity of Polycomb Repressive Complex 2. Mol Cell 70, 435–448 e435, doi:10.1016/j.molcel.2018.03.019 (2018).

23. O’Carroll, D. et al. The polycomb-group gene Ezh2 is required for early mouse development. Mol Cell Biol 21, 4330–4336, doi:10.1128/MCB.21.13.4330-4336.2001 (2001).

24. Ezhkova, E. et al. EZH1 and EZH2 cogovern histone H3K27 trimethylation and are essential for hair follicle homeostasis and wound repair. Genes Dev 25, 485–498, doi:10.1101/gad.2019811 (2011).

25. Volkel, P. et al. Ezh1 arises from Ezh2 gene duplication but its function is not required for zebrafish development. Sci Rep 9, 4319, doi:10.1038/s41598-019-40738-9 (2019).

26. Stojic, L. et al. Chromatin regulated interchange between polycomb repressive complex 2 (PRC2)-Ezh2 and PRC2-Ezh1 complexes controls myogenin activation in skeletal muscle cells. Epigenetics Chromatin 4, 16, doi:10.1186/1756-8935-4-16 (2011).

27. Bodega, B. et al. A cytosolic Ezh1 isoform modulates a PRC2-Ezh1 epigenetic adaptive response in postmitotic cells. Nat Struct Mol Biol 24, 444–452, doi:10.1038/nsmb.3392 (2017).

28. Mousavi, K., Zare, H., Wang, A. H. & Sartorelli, V. Polycomb protein Ezh1 promotes RNA polymerase II elongation. Mol Cell 45, 255–262, doi:10.1016/j.molcel.2011.11.019 (2012).

29. Ai, S. et al. Divergent Requirements for EZH1 in Heart Development Versus Regeneration. Circ Res 121, 106–112, doi:10.1161/CIRCRESAHA.117.311212 (2017).

30. Vo, L. T. et al. Regulation of embryonic haematopoietic multipotency by EZH1. Nature 553, 506–510, doi:10.1038/nature25435 (2018).

31. Xu, J. et al. Developmental control of polycomb subunit composition by GATA factors mediates a switch to non-canonical functions. Mol Cell 57, 304–316, doi:10.1016/j.molcel.2014.12.009 (2015).

32. von Schimmelmann, M. et al. Polycomb repressive complex 2 (PRC2) silences genes responsible for neurodegeneration. Nat Neurosci 19, 1321–1330, doi:10.1038/nn.4360 (2016).

33. Henriquez, B. et al. Ezh1 and Ezh2 differentially regulate PSD-95 gene transcription in developing hippocampal neurons. Mol Cell Neurosci 57, 130–143, doi:10.1016/j.mcn.2013.07.012 (2013).

34. Sobreira, N., Schiettecatte, F., Valle, D. & Hamosh, A. GeneMatcher: a matching tool for connecting investigators with an interest in the same gene. Hum Mutat 36, 928–930, doi:10.1002/humu.22844 (2015).

35. Deciphering Developmental Disorders, S. Large-scale discovery of novel genetic causes of developmental disorders. Nature 519, 223–228, doi:10.1038/nature14135 (2015).

36. Investigators, G. P. P. et al. 100,000 Genomes Pilot on Rare-Disease Diagnosis in Health Care - Preliminary Report. N Engl J Med 385, 1868–1880, doi:10.1056/NEJMoa2035790 (2021).

37. Karczewski, K. J. et al. The mutational constraint spectrum quantified from variation in 141,456 humans. Nature 581, 434–443, doi:10.1038/s41586-020-2308-7 (2020).

38. Grau, D. et al. Structures of monomeric and dimeric PRC2:EZH1 reveal flexible modules involved in chromatin compaction. Nat Commun 12, 714, doi:10.1038/s41467-020-20775-z (2021).

39. Justin, N. et al. Structural basis of oncogenic histone H3K27M inhibition of human polycomb repressive complex 2. Nat Commun 7, 11316, doi:10.1038/ncomms11316 (2016).

40. McCabe, M. T. et al. Mutation of A677 in histone methyltransferase EZH2 in human B-cell lymphoma promotes hypertrimethylation of histone H3 on lysine 27 (H3K27). Proc Natl Acad Sci U S A 109, 2989–2994, doi:10.1073/pnas.1116418109 (2012).

41. McCabe, M. T. et al. EZH2 inhibition as a therapeutic strategy for lymphoma with EZH2- activating mutations. Nature 492, 108–112, doi:10.1038/nature11606 (2012).

42. Yamagishi, M. et al. Targeting Excessive EZH1 and EZH2 Activities for Abnormal Histone Methylation and Transcription Network in Malignant Lymphomas. Cell Rep 29, 2321–2337 e2327, doi:10.1016/j.celrep.2019.10.083 (2019).

43. Stessman, H. A. et al. Targeted sequencing identifies 91 neurodevelopmental-disorder risk genes with autism and developmental-disability biases. Nat Genet 49, 515–526, doi:10.1038/ng.3792 (2017).

44. Li, M. et al. Integrative functional genomic analysis of human brain development and neuropsychiatric risks. Science 362, doi:10.1126/science.aat7615 (2018).

45. Chailangkarn, T. et al. A human neurodevelopmental model for Williams syndrome. Nature 536, 338–343, doi:10.1038/nature19067 (2016).

46. Paulsen, B. et al. Autism genes converge on asynchronous development of shared neuron classes. Nature 602, 268–273, doi:10.1038/s41586-021-04358-6 (2022).

47. Birey, F. et al. Dissecting the molecular basis of human interneuron migration in forebrain assembloids from Timothy syndrome. Cell Stem Cell 29, 248–264 e247, doi:10.1016/j.stem.2021.11.011 (2022).

48. Chambers, S. M. et al. Highly efficient neural conversion of human ES and iPS cells by dual inhibition of SMAD signaling. Nat Biotechnol 27, 275–280, doi:10.1038/nbt.1529 (2009).

49. Qian, X. et al. Sliced Human Cortical Organoids for Modeling Distinct Cortical Layer Formation. Cell Stem Cell 26, 766–781 e769, doi:10.1016/j.stem.2020.02.002 (2020).

50. Hu, H. et al. Genetics of intellectual disability in consanguineous families. Mol Psychiatry 24, 1027–1039, doi:10.1038/s41380-017-0012-2 (2019).

51. Jung, C. K. et al. Clinical utility of EZH1 mutations in the diagnosis of follicular-patterned thyroid tumors. Hum Pathol 81, 9–17, doi:10.1016/j.humpath.2018.04.018 (2018).

52. Calebiro, D. et al. Recurrent EZH1 mutations are a second hit in autonomous thyroid adenomas. J Clin Invest 126, 3383–3388, doi:10.1172/JCI84894 (2016).

53. Hu, Y. et al. Integrated Whole-Exome and Transcriptome Sequencing of Sporadic Parathyroid Adenoma. Front Endocrinol (Lausanne) 12, 631680, doi:10.3389/fendo.2021.631680 (2021).

54. Morin, R. D. et al. Somatic mutations altering EZH2 (Tyr641) in follicular and diffuse large B-cell lymphomas of germinal-center origin. Nat Genet 42, 181–185, doi:10.1038/ng.518 (2010).

55. Hodis, E. et al. A landscape of driver mutations in melanoma. Cell 150, 251–263, doi:10.1016/j.cell.2012.06.024 (2012).

56. Gibson, W. T. et al. Mutations in EZH2 cause Weaver syndrome. Am J Hum Genet 90, 110–118, doi:10.1016/j.ajhg.2011.11.018 (2012).

57. Hirabayashi, Y. et al. Polycomb limits the neurogenic competence of neural precursor cells to promote astrogenic fate transition. Neuron 63, 600–613, doi:10.1016/j.neuron.2009.08.021 (2009).

58. Akizu, N., Estaras, C., Guerrero, L., Marti, E. & Martinez-Balbas, M. A. H3K27me3 regulates BMP activity in developing spinal cord. Development 137, 2915–2925, doi:10.1242/dev.049395 (2010).

59. Pereira, J. D. et al. Ezh2, the histone methyltransferase of PRC2, regulates the balance between self-renewal and differentiation in the cerebral cortex. Proc Natl Acad Sci U S A 107, 15957–15962, doi:10.1073/pnas.1002530107 (2010).

60. Guy, J., Gan, J., Selfridge, J., Cobb, S. & Bird, A. Reversal of neurological defects in a mouse model of Rett syndrome. Science 315, 1143–1147, doi:10.1126/science.1138389 (2007).

61. Benjamin, J. S. et al. A ketogenic diet rescues hippocampal memory defects in a mouse model of Kabuki syndrome. Proc Natl Acad Sci U S A 114, 125–130, doi:10.1073/pnas.1611431114 (2017).

62. Meng, L. et al. Towards a therapy for Angelman syndrome by targeting a long non-coding RNA. Nature 518, 409–412, doi:10.1038/nature13975 (2015).

63. Mencacci, N. E. et al. De Novo Mutations in PDE10A Cause Childhood-Onset Chorea with Bilateral Striatal Lesions. Am J Hum Genet 98, 763–771, doi:10.1016/j.ajhg.2016.02.015 (2016).

64. Anazi, S. et al. Clinical genomics expands the morbid genome of intellectual disability and offers a high diagnostic yield. Mol Psychiatry 22, 615–624, doi:10.1038/mp.2016.113 (2017).

65. Li, D. et al. Pathogenic variants in SMARCA5, a chromatin remodeler, cause a range of syndromic neurodevelopmental features. Sci Adv 7, doi:10.1126/sciadv.abf2066 (2021).

66. den Dunnen, J. T. et al. HGVS Recommendations for the Description of Sequence Variants: 2016 Update. Hum Mutat 37, 564–569, doi:10.1002/humu.22981 (2016).

67. Wiel, L. et al. MetaDome: Pathogenicity analysis of genetic variants through aggregation of homologous human protein domains. Hum Mutat 40, 1030–1038, doi:10.1002/humu.23798 (2019).

68. Kojima, S., Vignjevic, D. & Borisy, G. G. Improved silencing vector co-expressing GFP and small hairpin RNA. Biotechniques 36, 74–79, doi:10.2144/04361ST02 (2004).

69. Langmead, B. & Salzberg, S. L. Fast gapped-read alignment with Bowtie 2. Nat Methods 9, 357–359, doi:10.1038/nmeth.1923 (2012).

70. Zhang, Y. et al. Model-based analysis of ChIP-Seq (MACS). Genome Biol 9, R137, doi:10.1186/gb-2008-9-9-r137 (2008).

71. Ramirez, F. et al. deepTools2: a next generation web server for deep-sequencing data analysis. Nucleic Acids Res 44, W160–165, doi:10.1093/nar/gkw257 (2016).

